# Heart Failure Prediction & Risk Stratification using Machine Learning

**DOI:** 10.64898/2026.04.03.26350139

**Authors:** Sher Ali, Mary Ann Leavitt, Waseem Asghar

**Affiliations:** Asghar-Lab, Micro and Nanotechnology in Medicine, College of Engineering and Computer Science, Boca Raton, FL 33431, USA; Department of Electrical Engineering and Computer Science, Florida Atlantic University, Boca Raton, FL 33431, USA; Christine E. Lynn College of Nursing, Florida Atlantic University, Boca Raton, FL, USA; Department of Biomedical Engineering (Affiliate Appointment), Florida Atlantic University, Boca Raton, FL 33431, USA

**Author notes:** Author to whom correspondence should be addressed: Waseem Asghar, Ph.D. 777 Glades Road, EE 435, Boca Raton, FL 33431-0991, USA.

## Abstract

Heart failure (HF) is one of the most prevalent causes of morbidity, mortality, and healthcare expenditures, with approximately 6.7 million adults in the U.S. suffering from this condition and contributing to hundreds of thousands of deaths annually. Early diagnosis of high-risk individuals has been a challenge, as the HF-specific symptoms are often ignored or misinterpreted as normal aging, stress, or minor illnesses, leading to delayed diagnosis. We trained, tested, and evaluated several models, including logistic regression, SVM, KNN, random forest, XGBoost, MLP, and a custom stacked ensemble using stratified 5-fold CV and 70/30 hold-out splits for HF prediction on routinely available electronic medical record (EMR) data of the All of Us Research Program. This group consisted of 37,070 adults (13,577 HF; 23,493 non-HF). The predictors included readily available demographics, vital signs, conditions, and laboratory results. Preprocessing steps included IQR-winsorization, median imputation, one-hot encoding, and QuantileTransformer. The stacked model obtained ROC-AUC 0.927, PR-AUC 0.895, and accuracy 0.856 in the test set. To support real-world deployment, we calibrated predicted probabilities and adjusted them to a realistic population prevalence, yielding interpretable probability estimates and clear stratification of individuals into clinically actionable risk tiers. SHAP analysis identified the most influential features, namely, atrial fibrillation, age, hypertensive disorder, sodium, and deprivation index, as the top 5 features impacting the model’s prediction. A secondary multiclass experiment (No-HF, HF with reduced ejection fraction, and HF with preserved ejection fraction) was performed, achieving lower discrimination results (macro-AUC ∼0.87) and a lower per-class precision/recall, presumably due to label noise, class imbalance, and overlapping phenotypes. We have demonstrated that a carefully calibrated stacked ensemble on the combination of readily available EMR variables can achieve strong discrimination on HF, making it an effective tool for an AI clinical decision support system (AI-CDSS) in population screening and proactive care pathways.

## Introduction

The United States alone has an estimated 6.7 million adults with HF. HF was reported on approximately 452,000 death certificates in 2023, making this condition a major cause of mortality in general ^1^. HF is estimated at 64 million individuals globally and is increasing with aging populations and improved treatment of HF ^2,3^, Widespread adoption of guideline-directed medical therapy allows patients to live longer but they still may experience a reduced quality of life. Predictive models in HF can improve patient outcomes by enhancing early detection, increasing person-centered treatment, and proactive, risk-based management ^4^. Electronic medical records (EMRs) provide a practical foundation for scalable prediction, as they comprise routinely collected data including demographics, vital signs, lab reports, and comorbidities documented as part of routine care. Recent investigations demonstrate that Machine Learning (ML) models applied to longitudinal EMR sequences can predict HF onset earlier than conventional statistical models, especially when temporal dynamics are represented by recurrent or attention-based neural networks ^5–8^. Yet, these models tend to involve dense and longitudinal measurements and custom pipelines that are difficult to implement uniformly in various clinical settings. This creates a need for interpretable, calibrated risk models built from commonly available EMR features that are operationalizable at scale.

In this study, we developed and evaluated EMR-based ML models to predict HF using data from the National Institutes of Health All of Us Research Program. This national patient database contains a rich collection of electronic health records, survey information, and physical measurements ^9,10^ We restricted our predictors to routinely collected variables such as demographics (age, sex at birth), vital signs and measurements (blood pressure, body mass index (BMI)), standard laboratory values (sodium, potassium, glucose, creatinine, albumin, hemoglobin, lipids), prevalent comorbidities (hypertension, atrial fibrillation, anemia, obesity), and lifestyle and social determinants (smoking history, neighborhood-level l deprivation index – also known as poverty index). We deliberately excluded specialized imaging, genomic data, or advanced biomarkers, focusing on features commonly available in standard clinical practice.

The work is presented to address a practical literature gap. Although high-performing temporal models are very accurate, they do not generalize well in cases where the dataset is imbalanced, and data density is low. Our study demonstrates that a well-calibrated, interpretable ensemble model using EMR variables can provide near state-of-the-art discrimination, thus supporting a low-cost AI-CDSS framework that can assist in population screening and proactive intervention for HF diagnosis. For HF subtypes, we present future work: a hierarchical strategy (identify HF first, followed by subtype HF cases), inclusion of longitudinal trends and echocardiographic values where possible. Our study reports both discrimination and model calibration, the latter being essential to decision support and often underreported in clinical ML ^11–13^. Third, we offer post-hoc explainability via SHAP analysis ^14^, providing a framework to understand individual feature contribution to the model’s prediction.

### Literature Review

The relentless progress in the area of artificial intelligence (AI) and ML has revolutionized the world of healthcare, providing it with opportunities to support clinical decisions based on data (diagnosis and prognosis). ML in cardiology has demonstrated specific potential in early prediction and risk stratification of HF ^15^. Despite this progress, the early detection of HF remains challenging due to the disease’s multifactorial nature, heterogeneity, and dependence on high-cost diagnostic modalities ^16^.

#### A. Imaging-Based Approaches

Much literature has been devoted to deep learning (DL) approaches in medical imaging. Matsumoto et al. ^17^ developed convolutional neural networks (CNNs) trained on chest X-rays (952 images) able to distinguish HF patients and controls with 82% accuracy, sensitivity of 75%, and specificity of 94.4%. Such results suggest that imaging-based DL systems may be useful in early HF detection, but they are constrained by their reliance on high cost of modalities.

Additionally, Choi et al. ^18^ presented algorithms that combined the use of imaging, including the CART-based system incorporating echocardiographic variables (LVEF, LAVI, and tricuspid-regurgitation velocity) to determine phenotypes of HF. Their model obtained 100% accuracy on HF with mid-range and reduced ejection fraction but lower performances on preserved ejection fraction (78.9%) and non-HF (80.5%). Despite its potential, the study’s cohort of 598 patients was relatively small, and features as determined by echocardiography need special equipment and operator skills.

#### B. Structured and EMR-Based Models

Beyond imaging, scholars have studied ML on structured clinical data mined out of electronic medical records (EMRs). Wu et al. ^19^ examined data from the Geisinger Clinic, which consisted of 179 variables, and used it to predict HF up to 6 months before clinical diagnosis with an AUC of 0.77 on 10-fold cross validation. Their model was based on logistic regression and boosting algorithms but employed an under-sampling technique, making their class size 536 each, and the cohort consisted mostly of older adults (50-79 years), limiting their study. Another study by Adler et al. ^20^ constructed a boosted decision tree, trained on 5822 HF patients, and combined a series of 8 laboratory and hemodynamic variables (e.g., diastolic blood pressure, creatinine, blood urea nitrogen, hemoglobin, albumin) for mortality prediction. The study achieved a risk score with an AUC of 0.88 with external validation on two separate HF datasets, attaining AUCs of 0.84 and 0.81, respectively, demonstrating that tabular EMR data can yield strong predictive power. However, the study was limited to using temporal data, and the dataset used for training was taken from a single center in San Diego, incurring demographic bias.

Ishaq et al. ^21^ recently reported the data-imbalance correction approach based on the Synthetic Minority Oversampling Technique (SMOTE), paired with the reliable feature selection method to enhance HF survival prediction. The study used a dataset of 299 heart failure survivors and aimed to find relevant features associated with HF patients’ survival. Their approach showed significantly enhanced sensitivity, and F1-score compared with traditional models, emphasizing the need to take the class imbalance issue in clinical datasets into consideration. They achieved an accuracy of 92.6% and an F1 score, precision, and recall of 93%. Other complementary studies with interpretable ML, including Wang (2021) ^22^, included SHAP to help explain 3-year mortality risk in coronary-heart-disease-related HF, promoting clinician trust and transparency.

#### C. Limitations in Current Studies

Although studies discussed in previous sections show good results, a number of limitations impede clinical translation.

1. Limitations of the datasets: The majority of the studies were based on single-center or geographically small cohorts, which limits the model extrapolation to other populations with other lifestyles or access to care.
2. Dependency on data type: Most of the algorithms use advanced imaging or intrusion techniques (e.g., CT, MRI, CardioMEMS, V-LAP) that are absent in primary-care or resource-constrained settings.
3. Interpretability and calibration of models: Other studies very rarely report probability calibration, or give interpretable descriptions of features, which are essential to regulatory and clinical acceptance.
4. Temporal dependency: There are several EHR-based models using longitudinal data. Sampling data across time is not always practical in real world especially in disease diagnosis applications.

#### D. Research Gap and Present Study

Examining the literature reviewed, it is evident that the development of HF-prediction studies has shifted to models being EMR-driven. However, model generalizability, model calibration, and applicability to clinical interpretation remain critical weaknesses in model development. The existing literature is limited to local datasets, expensive body imaging, or invasive monitoring technologies, limiting the ability of these methods to scale up to a population-wide screening. Moreover, evaluation scores in some studies are mainly based on accuracy or ROC-AUC, which are not indicative of the class-imbalanced data used in medical studies, e.g., HF prediction where the occurrence of positive cases is less frequent. In this case a model can have high ROC-AUC even with a low precision on the minority class, resulting in excessive false positives or missed true HF cases ^23^. To overcome this, our study emphasizes the Area Under the Precision–Recall Curve (PR-AUC), which is the metric that directly, and more accurately, represents the trade-off between precision (positive predictive value) and recall (sensitivity) in an imbalanced dataset setting ^23^.

## Methodology

We followed best practices for clinical prediction modelling, consisting of three crucial steps. First, for data preprocessing, we performed IQR-winsorization ^24^, median imputation, one-hot encoding ^25^ of categorical variables, and QuantileTransformer ^26^ normalization to stabilize model training across heterogeneous value ranges. Second, we benchmark a diverse range of algorithms (logistic regression, SVM, KNN, random forest, gradient-boosted trees, multilayer perceptron) and a stacked ensemble containing XGBoost, LightGBM, CatBoost, and MLP as base models and logistic regression as meta-learner, all followed by stratified 5-fold cross-validation and with a 70/30 hold-out split. Third, the predicted probabilities were calibrated via Platt scaling and prior probabilities were shifted to reflect real world HF prevalence rate for clinical reliable decision making.

Figure 1 presents an analytical workflow to predict incident HF and stratify patients into clinically meaningful risk tiers using routinely available EMR data. This pipeline begins with data curation and data preprocessing, in which heterogeneous patient records are acquired, cleaned, standardized, and encoded. This preprocessed dataset is then passed to ML models for training. The model output supports a binary classification for distinguishing HF. The same output probabilities are then calibrated and routed to the risk stratification module, where patients are categorized in risk deciles, later explained in the model calibration and risk stratification section.

**Figure 1:**
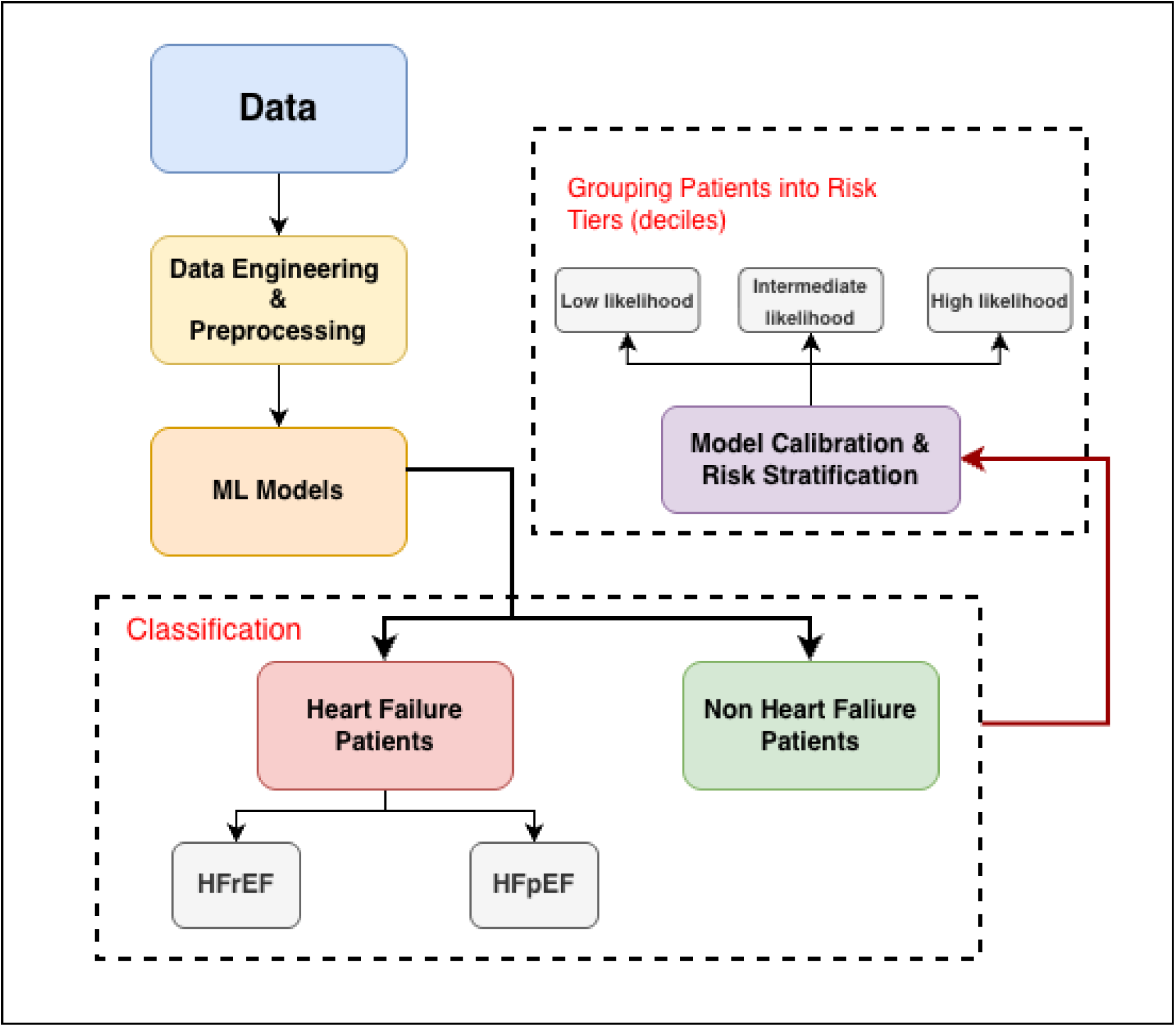
ML pipeline for HF prediction and risk stratification

### Ethics Approval and Consent to Participate

For this study we used the National Institutes of Health’s *All of Us* research program’s de-identified EMR dataset, a large U.S. biomedical database designed to promote data-driven health research across diverse populations. The access to this dataset can be obtained via Researcher Workbench, which uses a Passport Access Model. Users need to fulfill the compliance requirement to create a workspace and do their analysis, which is restricted to the Jupyter Notebook platform provided on their own website; no participant-level data can be downloaded or be published. The institution the user belongs to has to sign the Data Use and Registration Agreement (DURA), provided at https://www.researchallofus.org/register/. The *All of Us* IRB approved the *All of Us* protocol and material and obtained informed consent from all participants at the time of enrollment; hence, no additional consent was required for this secondary analysis.

### Dataset

We constructed a cohort of 37,070 adults, comprising 13,577 (36.6%) patients with a clinical diagnosis of HF and 23,493 (63.4%) non-HF controls, labels being derived using ICD9 and ICD10CM diagnostic codes and confirmed by the mapped standard vocabulary (SNOMED) from the structured EMR entries. It is important to note that the dataset composition is not based on the real-world prevalence of the disease, nor the total number of HF and non-HF patients present in the All of Us dataset but rather on the basis of data availability and applicability within the scope of study. Several parameters were set while deciding which patients’ entries should be included. For example, we excluded patients who were deceased, terminally ill, or had cancer.

Figure 2 depicts a descriptive analysis of the demographics of this cohort. Notably, in the HF category, patients were generally older, with ages clustering between 60 – 85 years and a distribution peaking around the mid-70s. Whereas, for non-HF patients, we observe a significantly smaller population of older patients with a peak at age 65. For each patient’s sex, we acquired their appointed sex at birth. For HF, sex distribution was approximately balanced between females and males (51% vs 49%), while the non-HF group showed a female predominance (68% vs 32%). These differences are consistent with the prior literature of epidemiologic evidence that HF prevalence increases with age and differs by sex ^2,27^.

**Figure 2:**
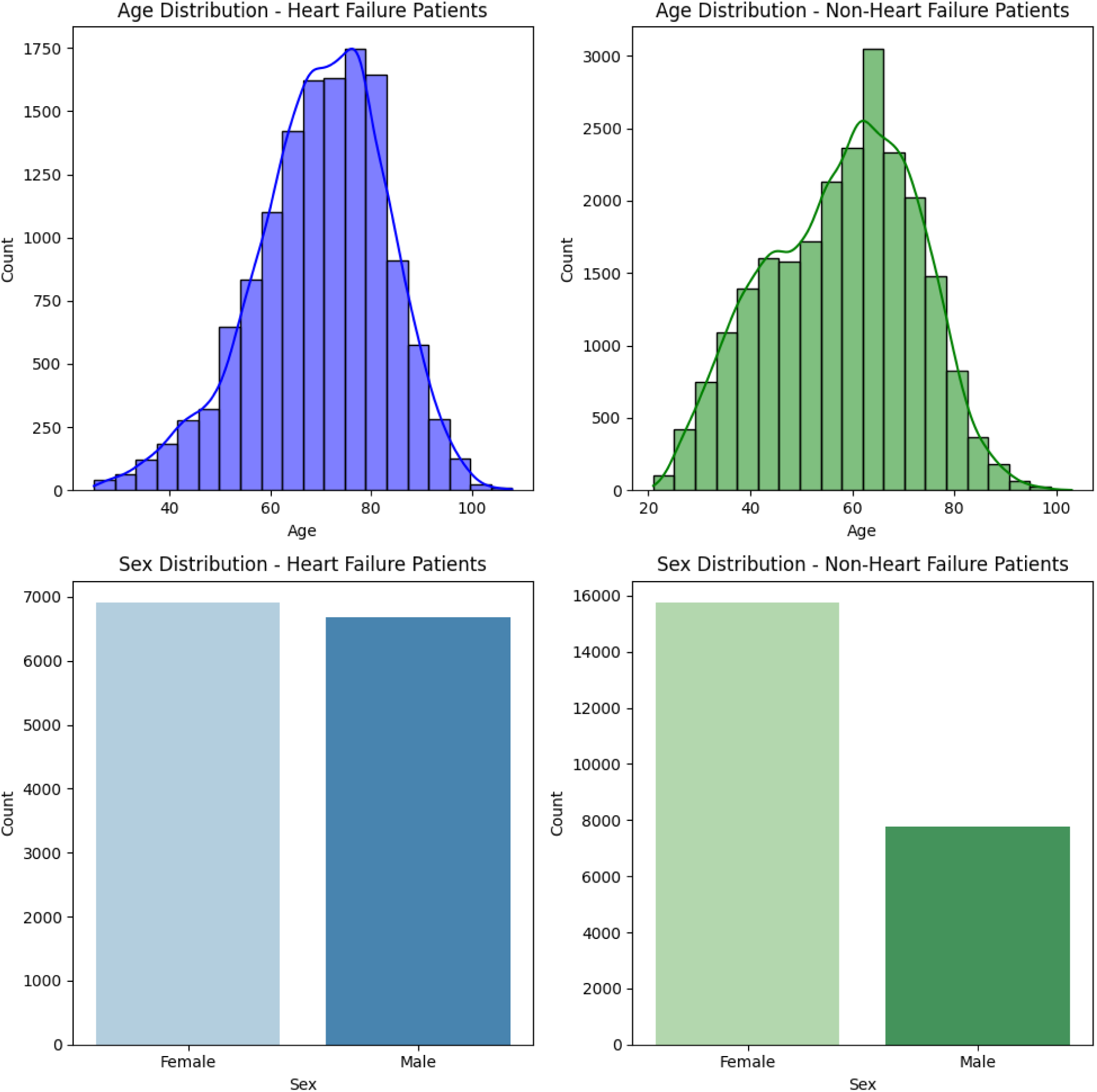
Demographic analysis of HF and non-HF patients

For the second part of the experiment, we categorized HF into subcategories. Each HF record contained a “standard_concept_name” entry corresponding to a clinical terminology for the HF type. These terms were mapped into three parent diagnostic categories based on left ventricular ejection fraction (LVEF): Heart failure with reduced ejection fraction (HFrEF: 40% or less LVEF), heart failure with preserved ejection fraction (HFpEF: greater than 40% LVEF) ^3^, and non-HF patients. The categorization, as per the literature, was made by identifying the keywords “systolic” for HFrEF and “diastolic” for HFpEF according to the guideline ^28^. Unfortunately, the database contained several heart failure entries without either “systolic” or “diastolic” identifiers. These entries were excluded from the analysis. This brought the HF category total count to 9257 patients (5426 HFrEF & 3831 HFpEF). The classification logic and the corresponding terminology groups are summarized in table 1.

**Table 1:** Mapping of HF terminologies into subcategories.

| Parent Category | Representative Terms (standard_concept_name) |
| --- | --- |
| HFrEF | “systolic heart failure,”<br>“chronic systolic heart failure,”<br>“acute systolic heart failure,”<br>“acute on chronic systolic heart failure,”<br>“heart failure with reduced ejection fraction” |
| HFpEF | “diastolic heart failure,”<br>“chronic diastolic heart failure,”<br>“acute diastolic heart failure,”<br>“acute on chronic diastolic heart failure,”<br>“heart failure with normal ejection fraction,”<br>“heart failure with preserved ejection fraction” |

These subcategory distributions are visualized in Figure 3, showing that the HFrEF group constituted the largest HF subtype and HFpEF the smallest, consistent with national epidemiologic data ^29^. The inclusion of standard-concept HF phenotypes and non-HF controls enables both binary and multiclass training configurations, allowing the model to determine (i) the presence/absence of HF and (ii) subtype differentiation (HFrEF & HFpEF).

**Figure 3:**
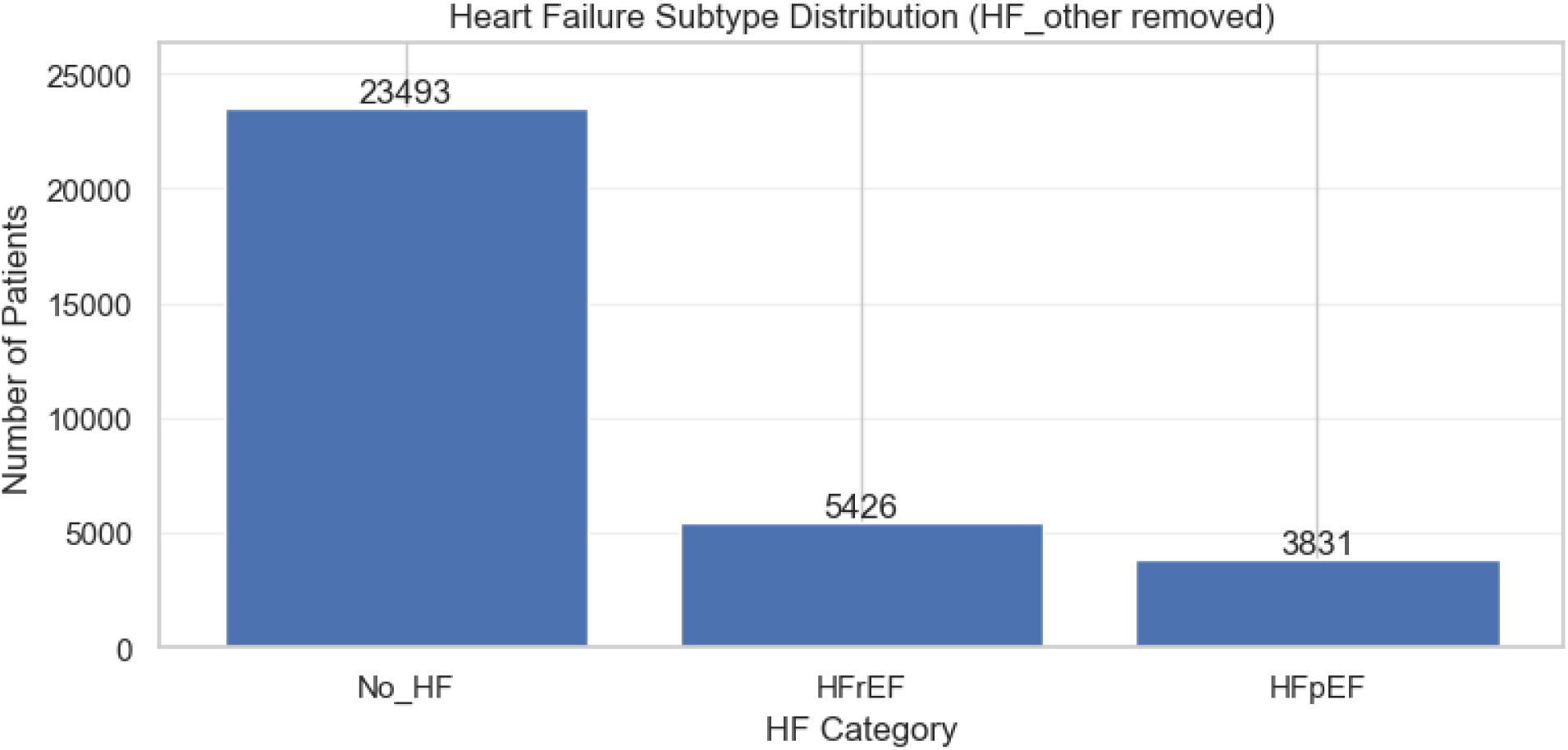
Distribution of HF subcategories within the dataset (No HF, HFrEF, and HFpEF).

### Feature Selection

This study utilized 18 attributes for HF evaluation categorized into four primary domains: Measurements, Conditions, Lifestyle, and Demographics, and they are displayed categorically in table 2. Feature Selection was guided primarily by their clinical relevance to HF in literature. However, three critical factors were taken into account, enabling the population-wide applicability and future incorporation of this study into an AI-CDSS.

1. Clinical relevance: features supported by cardiovascular evidence linking the parameters to HF onset, severity, or diagnosis.
2. Data completeness: ensuring high coverage across patient records in the cohort.
3. Accessibility: focusing essentially on routinely measured health records rather than expensive or inaccessible in primary-care or resource-constrained settings.

**Table 2:** feature set used for the prediction of the HF. The cohort includes four groups of categories: lab measurements, conditions, lifestyle, and demographics of the patients. Numerical features are marked with (#) and categorical features with (*).

| Measurements | Conditions | Lifestyle | Demographics |
| --- | --- | --- | --- |
| Albumin (m/v in Serum or Plasma) # | Anemia* | Smoked more than 100 cigarettes in a lifetime * | Age # |
| BMI # | Atrial Fibrillation * | Deprivation Index # | Sex at birth * |
| Creatinine (m/v in Serum or Plasma) # | Obesity * |  |  |
| Glucose (m/v in Serum or Plasma) # | Hypertensive disorder * |  |  |
| Hemoglobin (m/v in Serum or Plasma) # |  |  |  |
| Potassium (m/v in Serum or Plasma) # |  |  |  |
| Sodium (m/v in Serum or Plasma) # |  |  |  |
| Systolic Blood Pressure # |  |  |  |
| Diastolic Blood Pressure # |  |  |  |
| HDL Cholesterol (m/v in Serum or Plasma) |  |  |  |

Collectively, the selected variables cover a broad spectrum from systemic physiology to chronic comorbidities, which underlyingly relates to HF development. For example, just as creatinine and low albumin relate to renal impairment and nutritional depletion, BMI and hypertension are precursors for metabolic overload. Similarly, atrial fibrillation and anemia are well-established indicators of cardiac remodeling and reduced cardiac output ^30,31^. Literature also maps a direct relation between chronic HF and various comorbidities, including but not limited to anemia, significantly impacting the prognosis of patients with HF ^32^.

Likewise, a feature like the deprivation index (a measure of poverty) enables the model to account for socioeconomic gradients in HF risk, which in literature has proved to mediate disparities in cardiovascular outcomes ^33^ and is often ignored by researchers aiming to develop AI-CDSS for HF diagnosis. This multi-domain design for features allows our model to integrate several aspects, i.e., physiological, behavioral, and demographic factors, providing a comprehensive representation of each patient’s health profile while maintaining explainability for clinicians.

### Data Preprocessing

After feature selection, the final cohort had 18 features and 37070 patient entries. Naturally, not all these patients’ entries are populated. They have some level of missingness in the dataset; heterogeneous clinical distribution is often skewed, and as seen from the demographic’s distribution, our cohort faces a class imbalance. Hence, a proper preprocessing framework is adopted in this study following a rigorous and reproducible sequence, ensuring data consistency and statistical stability before data is fed to the ML models.

#### 1) Handling Missingness and Outliers

All continuous numerical variables were first examined for completeness and outlier presence. Missing entries were handled using median imputation, a central tendency estimator preserving the original feature distribution while preventing bias toward extreme values ^34^. The process includes mitigating the influence of measurement errors and rare anomalies by using interquartile range (IQR)-based winsorization. This technique caps extreme values at the 1.5xIQR threshold for both tails of the data distribution. Numerical variables like Age, BMI, Albumin, Sodium, Potassium, Glucose, Creatinine, HDL, and Blood Pressure, etc. are processed using this method. Figure 4 illustrates the data distribution before transformation. Skewness can be seen for laboratory biomarkers such as glucose and creatinine, reflecting the heterogeneity of underlying disease severity among patients.

**Figure 4:**
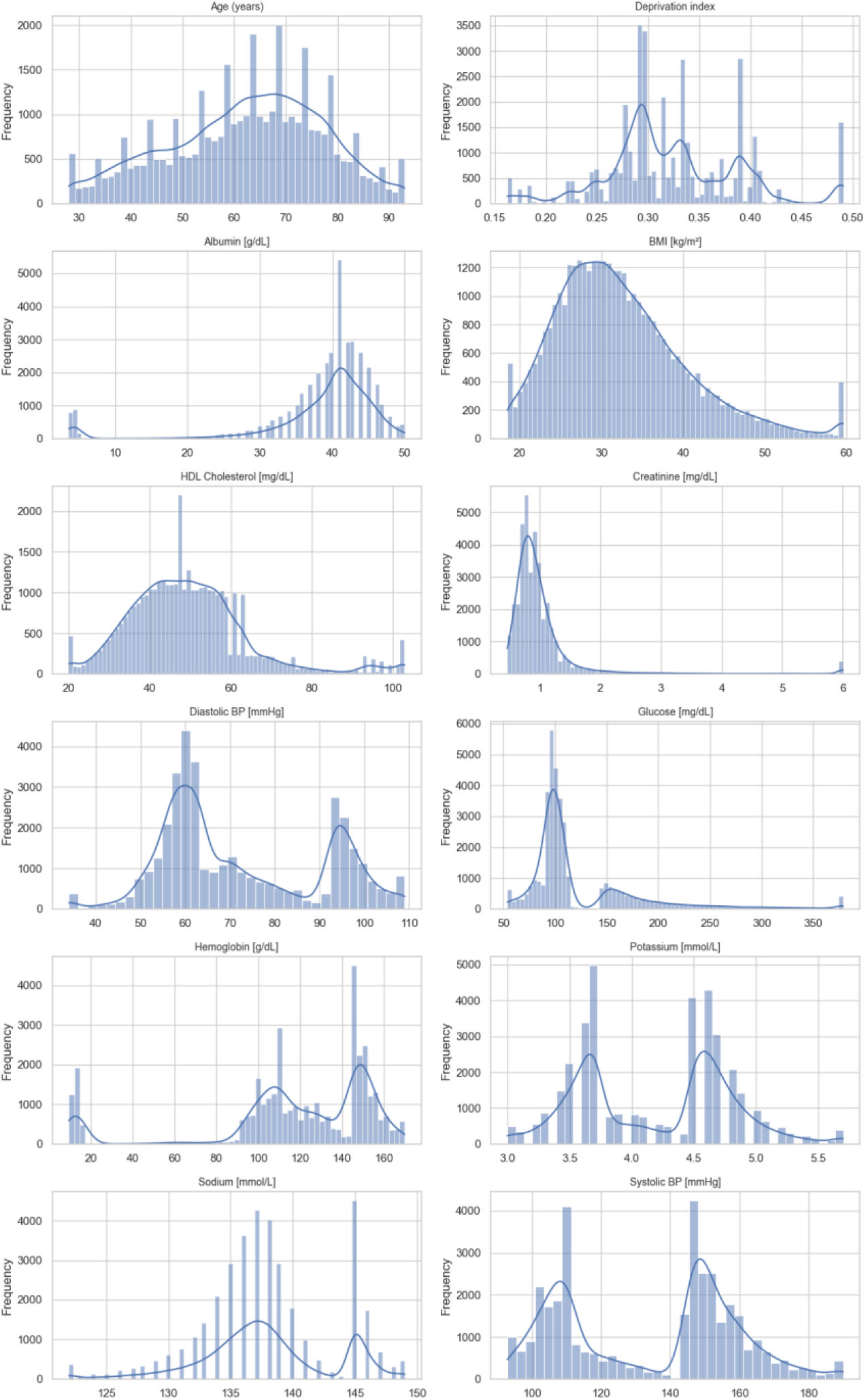
Raw distribution of continuous clinical variables before normalization

#### 2) Normalization and Variance Stabilization

As seen in figure 4, data distribution demonstrates a highly non-Gaussian nature of the EMR features. Hence, as many models are sensitive to feature scaling (e.g., Support Vector Machines, Logistic Regression, and Neural Networks) ^35^, we employed Quantile Transformer to each continuous variable to achieve approximate normality. This transformation maps the empirical cumulative distribution of each variable to a standard normal distribution (*N* (0,1)) while preserving relative ranking across patients ^26^.

Figure 5 represents the transformed variables, showing a close alignment with the theoretical Gaussian density, with uniform variance across all clinical features. The corresponding Q–Q plots (Figure 6) confirm strong linearity against the *N* (0,1) quantiles, indicating successful distributional normalization ^36^.

**Figure 5:**
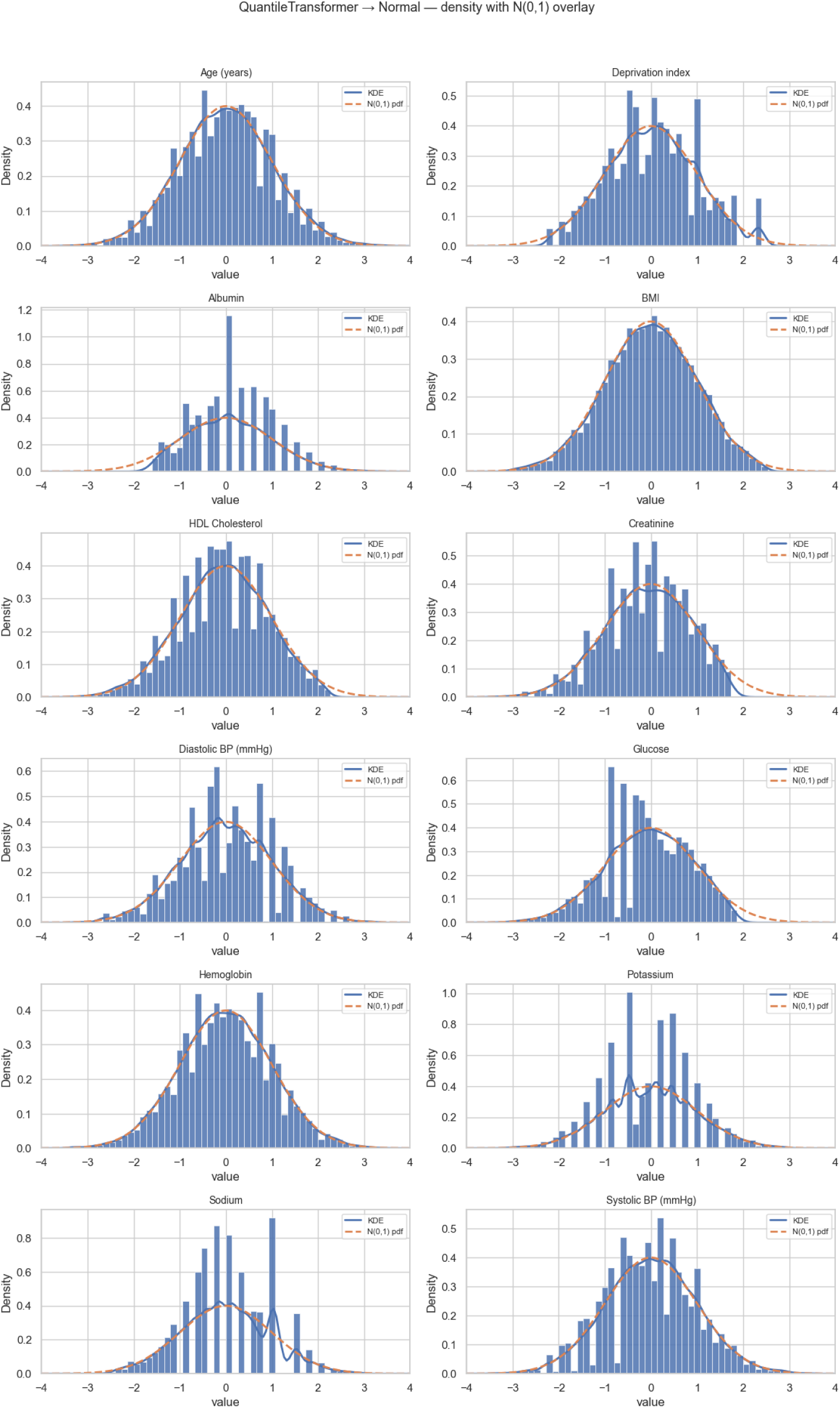
Post-transformation density plots after applying QuantileTransformer and IQR-based winsorization, showing alignment with normal distribution

**Figure 6:**
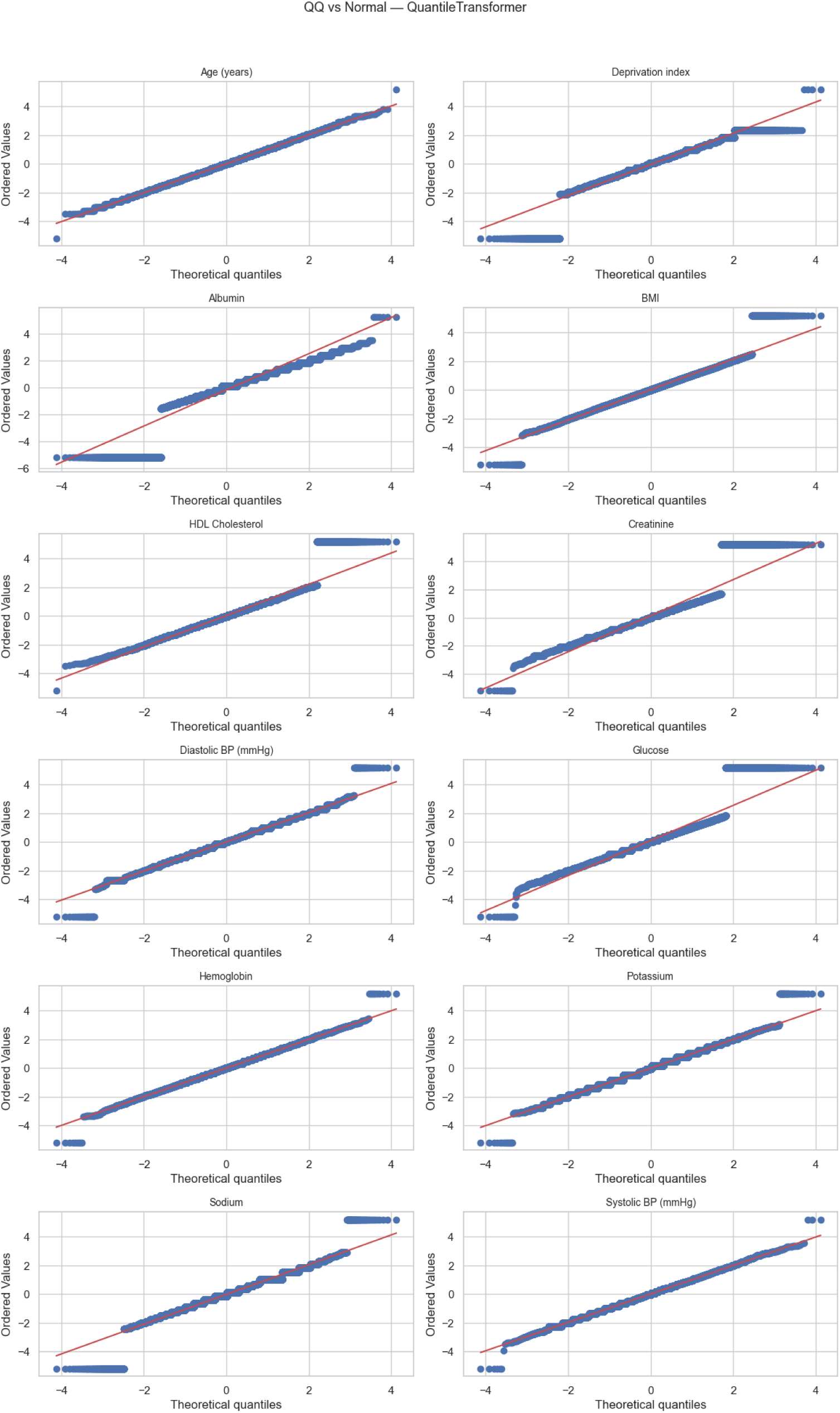
Q-Q plots verifying normality of transformed variables across all measurement features.

#### 3) Encoding Categorical Variables

The categorical features such as Sex at birth, Hypertension, Atrial fibrillation, Smoking status, Obesity, and Anemia were converted using one-hot encoding. This allowed each binary indicator to represent the presence or absence of the condition, ensuring that categorical information could be interpreted consistently across ML models ^25^.

#### 4) Train–Test Partitioning and Cross-Validation

The dataset was partitioned using a 70/30 stratified train/test split. To prevent data leakage, all preprocessing steps, including imputation, winsorization, and quantile transformation, were fit exclusively on the training data ^37^. Within the training set, we used 5-fold cross-validation, in each fold, the preprocessing step was fit to only the training portion and applied to the fold’s validation portion to improve model performance and ensure consistent generalized performance across subsamples. The final model was then trained on the full training set and evaluated once on the held-out test set.

#### 5) Addressing Class Imbalance

Our cohort had a class imbalance: HF (13,577 cases) and non-HF (23,493 controls). To mitigate that, various strategies were employed, and a comparative analysis was performed on the empirical results of the model. These strategies included class reweighting, where we applied different weights to classes, prioritizing the minority class, and data resampling. Primary training relied on class weights being inversely proportional to class frequencies, ensuring balanced contribution of each group. Experiments were then conducted using the original distribution of classes, the dataset with SMOTE, and finally with random undersampling to observe how each technique affects predictive results ^38^.

This combination of reweighting and augmentation also helped achieve fair sensitivity–specificity trade-offs across subtypes while improving recall for minority HF classes, especially in the multiclass setting (HFrEF, HFpEF, non-HF). A complete discussion is presented in the Machine Learning section of the paper.

## Primary Experiment: Binary Class Heart Failure Classification

### Custom Stacked Ensemble Architecture

We developed a pipeline for a custom stacked ensemble model. The proposed stacked ensemble consists of four base learners: XGBoost, LightGBM, CatBoost, and MLP, and a meta learner: LR. Training followed a 5-fold stratified cross-validation approach, where out-of-fold (OOF) predictions from each base model were used as meta-features for LR training. A complete architecture is represented in figure 7 below:

**Figure 7:**
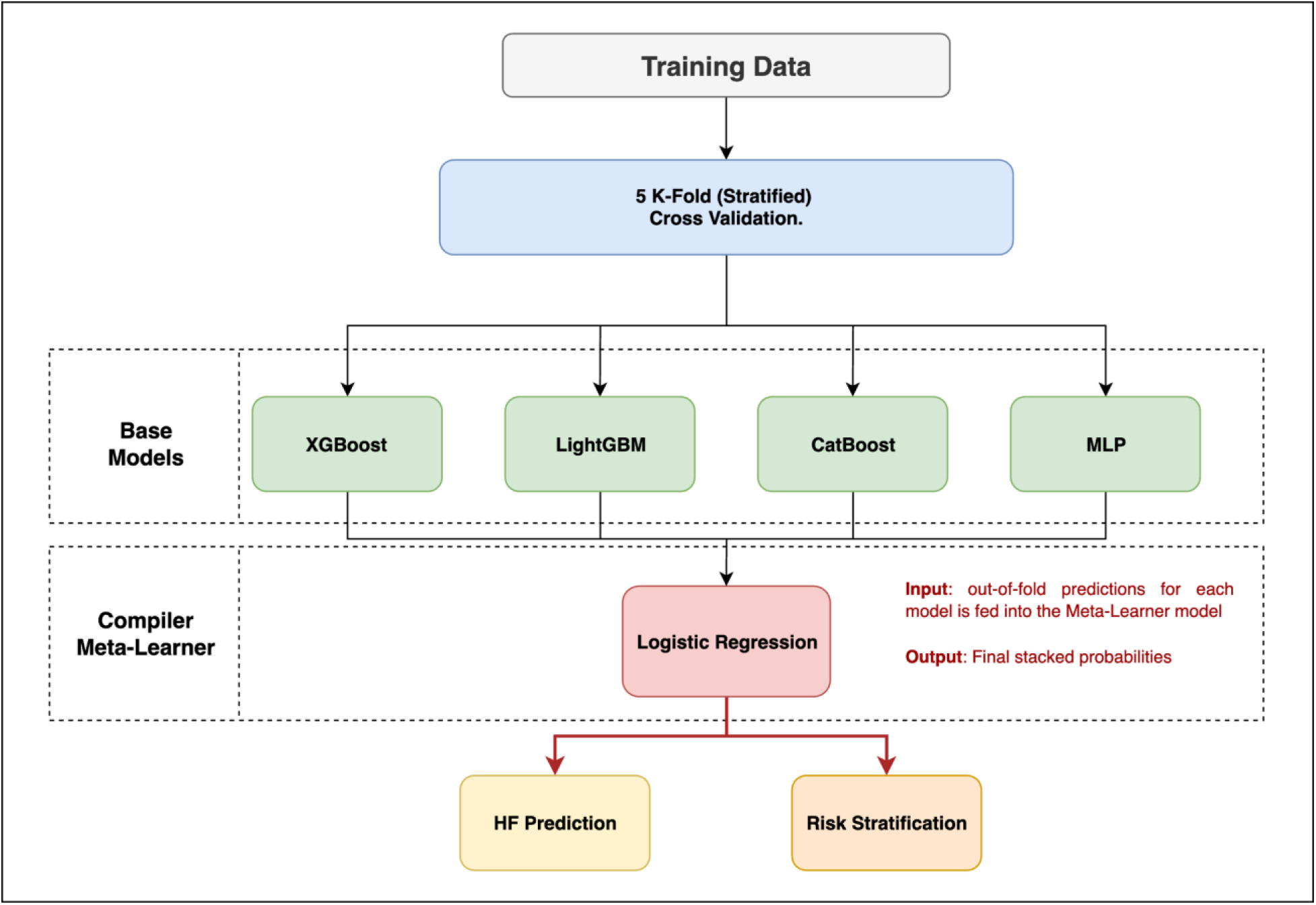
Stacked ensemble pipeline integrating tree-based and neural models via Logistic Regression meta-learner

Each model’s best hyperparameters were obtained via grid search, and the best threshold value was used for testing. Complete configuration and the rationale for choosing each base model and meta-model are presented in table 3.

**Table 3:**
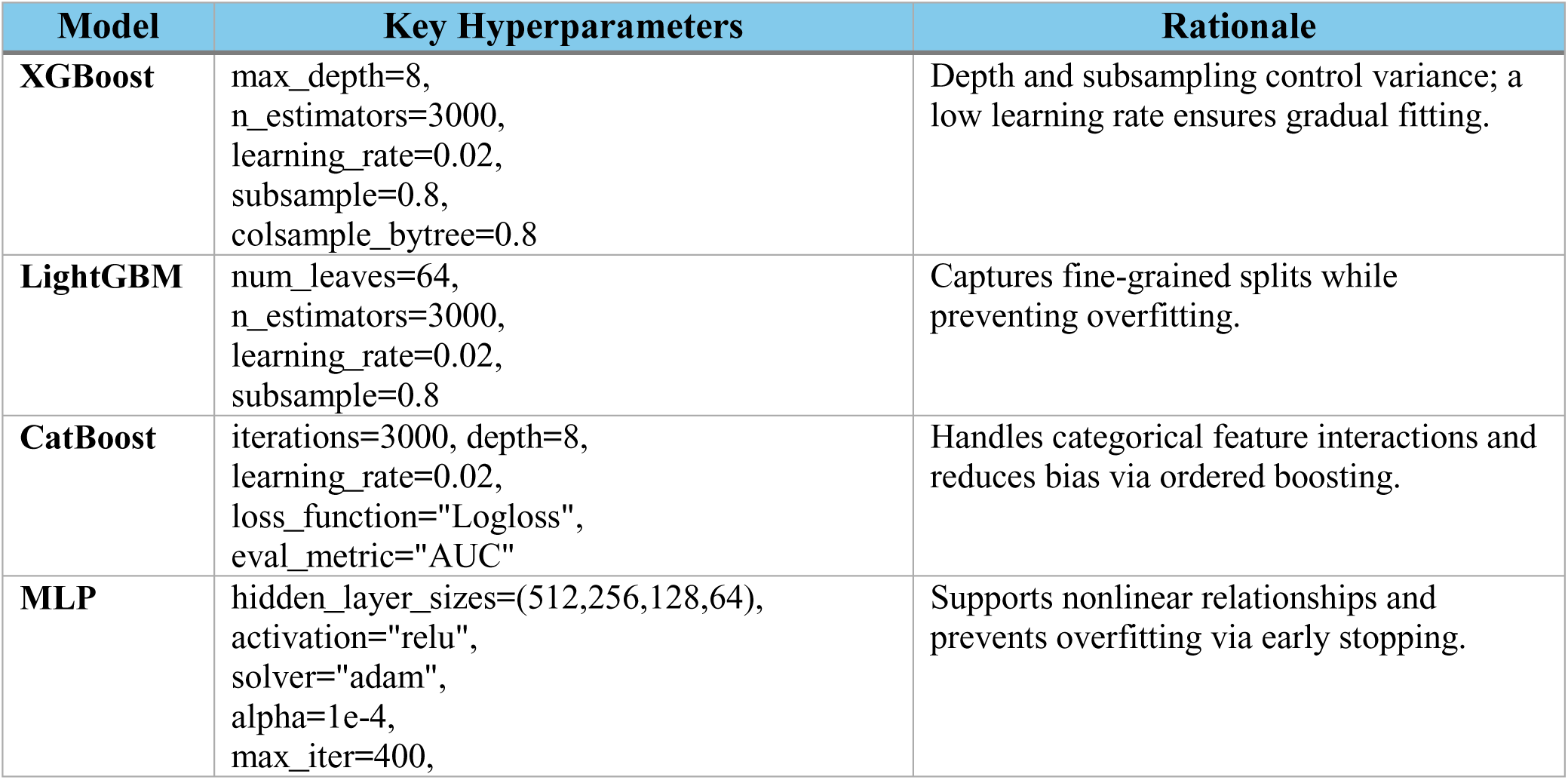
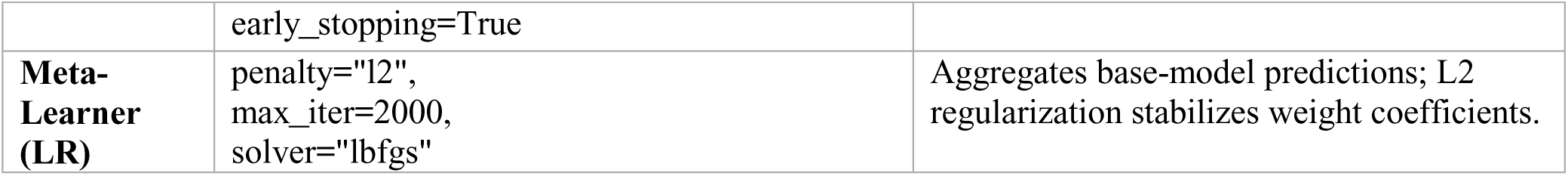
Model-specific hyperparameters and reasoning for choosing each base model.

| Model | Key Hyperparameters | Rationale |
| --- | --- | --- |
| **XGBoost** | max_depth=8,<br>n_estimators=3000,<br>learning_rate=0.02,<br>subsample=0.8,<br>colsample_bytree=0.8 | Depth and subsampling control variance; a low learning rate ensures gradual fitting. |
| **LightGBM** | num_leaves=64,<br>n_estimators=3000,<br>learning_rate=0.02,<br>subsample=0.8 | Captures fine-grained splits while preventing overfitting. |
| **CatBoost** | iterations=3000, depth=8,<br>learning_rate=0.02,<br>loss_function="Logloss",<br>eval_metric="AUC" | Handles categorical feature interactions and reduces bias via ordered boosting. |
| **MLP** | hidden_layer_sizes=(512,256,128,64),<br>activation="relu",<br>solver="adam",<br>alpha=1e-4,<br>max_iter=400, | Supports nonlinear relationships and prevents overfitting via early stopping. |
|  | early_stopping=True |  |
| **Meta-Learner (LR)** | penalty="l2",<br>max_iter=2000,<br>solver="lbfgs" | Aggregates base-model predictions; L2 regularization stabilizes weight coefficients. |

During training, class weights were incorporated into the meta-learner to address the imbalance between HF (positive) and non-HF (negative) cases. The base learners were trained under three distinct resampling schemes:

- Balanced (Undersampling): Randomly downsampling the majority class to match minority cases (HF: 13,577 vs. Non-HF: 13,577).
- SMOTE (Oversampling): Oversampling the HF class to equal the majority count (HF: 23,493 vs. Non-HF: 23,493).
- Unbalanced (Raw): Original dataset retained (HF: 13,577 vs. Non-HF: 23,493), with class weights applied during optimization.

Table 4 summarizes the results of each iteration.

**Table 4:** Stacked ensemble performance under different training strategies.

| SETUP (TRAINING STRATEGY) | ROC-AUC | PR-AUC | ACCURACY | PRECISION (NON-HF / HF) | RECALL (NON-HF / HF) | F1 (NON-HF / HF) | MACRO AVG (PREC / REC / F1) | WEIGHTED AVG (PREC / REC / F1) |
| --- | --- | --- | --- | --- | --- | --- | --- | --- |
| **BALANCED (UNDERSAMPLE)** | 0.924 | 0.891 | 0.838 | 0.908 / 0.742 | 0.829 / 0.854 | 0.866 / 0.795 | 0.825 / 0.842 / 0.831 | 0.847 / 0.838 / 0.840 |
| **SMOTE (OVERSAMPLE)** | 0.924 | 0.892 | 0.851 | 0.868 / 0.819 | 0.903 / 0.763 | 0.885 / 0.790 | 0.844 / 0.833 / 0.838 | 0.850 / 0.851 / 0.850 |
| **UNBALANCED (RAW DATA)** | **0.927** | **0.895** | **0.856** | **0.865 / 0.837** | **0.916 / 0.753** | **0.890 / 0.793** | **0.851 / 0.834 / 0.841** | **0.855 / 0.856 / 0.854** |

While undersampling and SMOTE yielded similar ROC-AUC (0.924), the unbalanced (raw) iteration slightly outperformed others, achieving ROC-AUC of 0.927, PR-AUC of 0.895, and accuracy of 0.856. This suggests that our original data distributions (Raw) combined with class weights achieved better overall results for the test set. To further visualize the evaluation of this model, ROC and PR-AUC curves and a confusion matrix of the unbalanced setup are demonstrated in figure 8.

**Figure 8:**
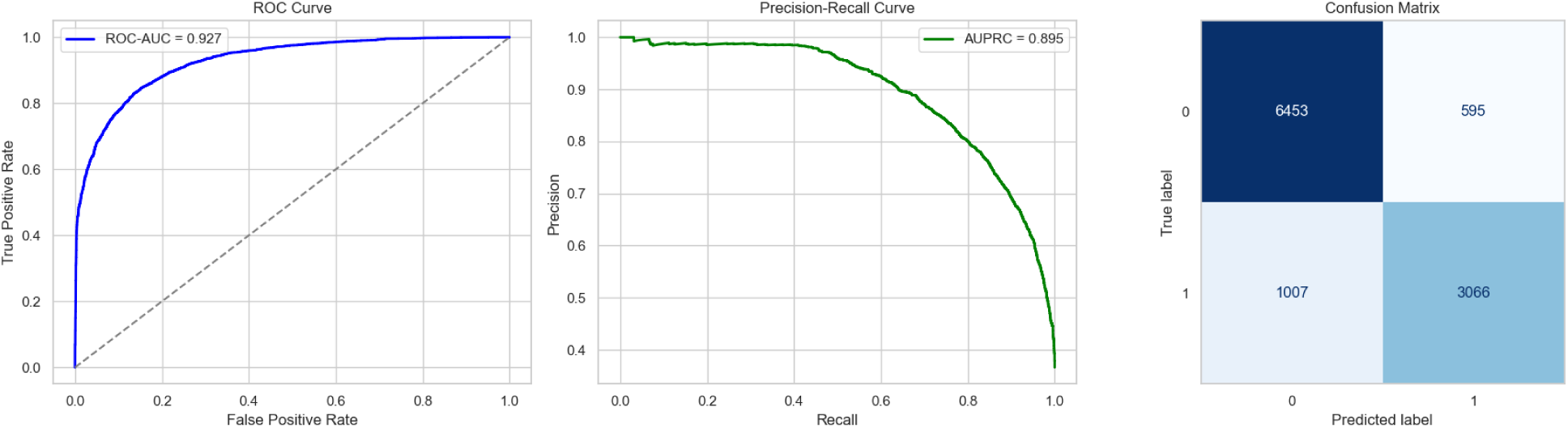
Evaluation plots of stacked ensemble(unbalanced) on test set (ROC-AUC, PR-AUC, and confusion matrix).

The ensemble achieved a smooth ROC curve and a high PR-AUC of 0.895, confirming strong predictive precision for the minority (HF) class, which is a crucial consideration in clinical risk modeling. The confusion matrix shows a true positive count of 3,066 and a true negative count of 6,453 with a small percentage of false negatives and false positives.

### Machine Learning Model Evaluation and Comparative Analysis

Further evaluations were done to benchmark on classical and deep learning models, including Logistic Regression (L1), Support Vector Machine (SVM) with RBF kernel, K-Nearest Neighbors (KNN), Random Forest (RF), XGBoost (XGB), and a Multi-Layer Perceptron (MLP) trained with 5-fold cross-validation, a 70/30 train-test split, and SMOTE. The same evaluation metrics were computed as done in the stacked ensemble model section, including ROC-AUC, PR-AUC, Precision/Recall/F1, calibration, and explainability assessments.

The results are compiled in table 5. XGBoost achieved the highest individual ROC-AUC (0.929) and PR-AUC (0.898), marginally outperforming the ensemble model (0.927 and 0.895, respectively), while it achieves lower accuracy and precision. Tree-based models dominated tabular EMR data due to their ability to capture non-linear feature interactions, while the linear and distance-based classifiers (LR, SVM, KNN) performed moderately well.

**Table 5:** ML models performances using 5-fold cross validation, SMOTE, and 70/30 train test compared to stacked ensemble model (unblanaced)

| *Model* | *Threshold* | *ROC-AUC* | *PR-AUC* | *Accuracy* | *Precision (macro)* | *Recall (macro)* | *F1-Score (macro)* |
| --- | --- | --- | --- | --- | --- | --- | --- |
| *Stacked Ensemble* | **0.50** | **0.927** | **0.895** | **0.856** | **0.851** | **0.834** | **0.841** |
| *XGBoost* | 0.51 | 0.929 | 0.898 | 0.853 | 0.840 | 0.849 | 0.844 |
| *Random Forest* | 0.45 | 0.921 | 0.889 | 0.851 | 0.840 | 0.840 | 0.840 |
| *SVM (RBF)* | 0.48 | 0.910 | 0.867 | 0.836 | 0.822 | 0.830 | 0.826 |
| *Logistic Regression (L1)* | 0.49 | 0.893 | 0.863 | 0.823 | 0.809 | 0.813 | 0.811 |
| *KNN* | 0.47 | 0.903 | 0.867 | 0.830 | 0.816 | 0.820 | 0.818 |
| *MLP* | 0.55 | 0.861 | 0.827 | 0.793 | 0.776 | 0.779 | 0.778 |

For analysis, the ROC curves for these traditional models were plotted on a single plot, revealing strong discriminative ability across models (Fig. 9).

**Figure 9:**
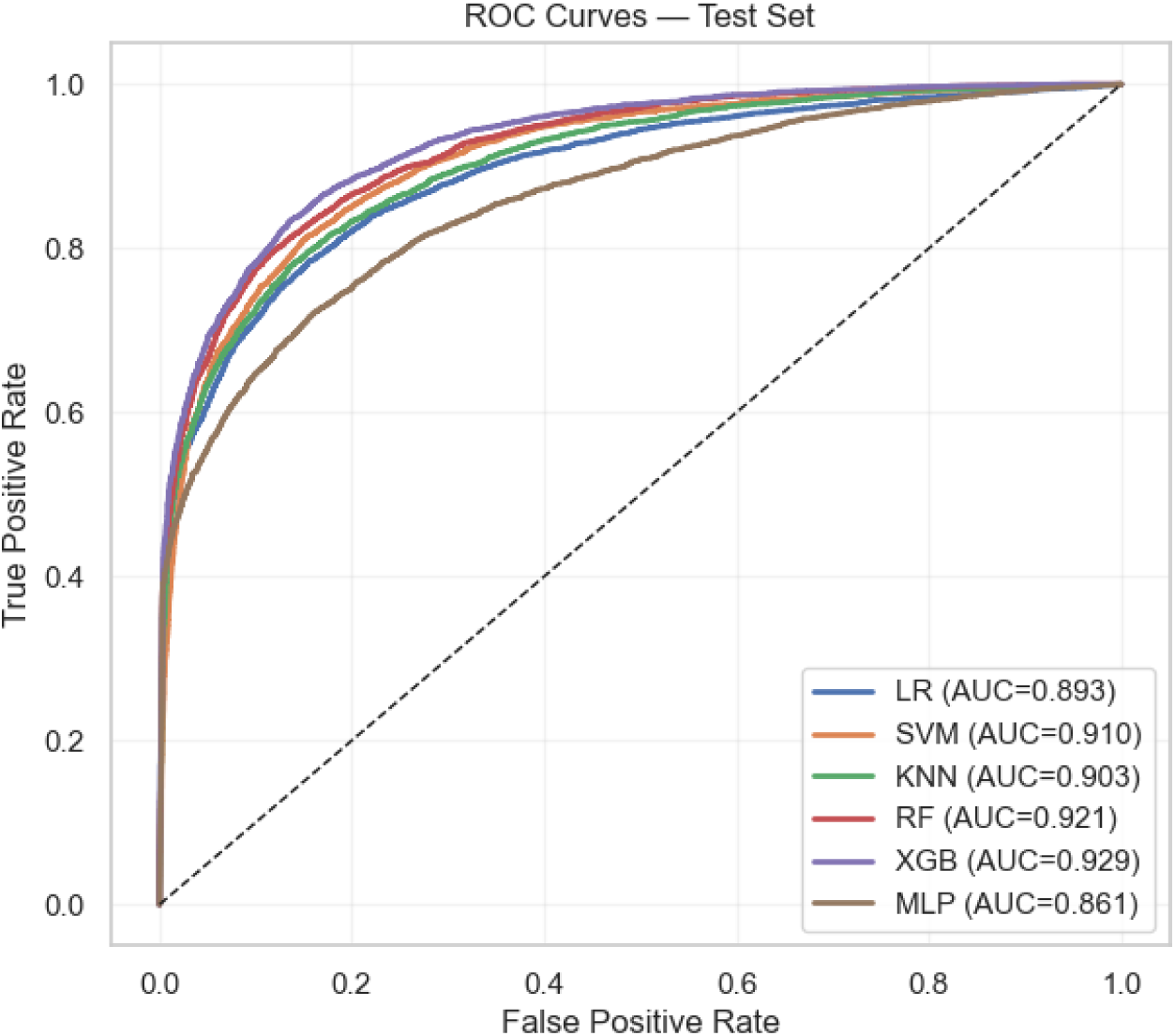
ROC-AUC graphs of the ML models

However, as our dataset is moderately imbalanced (≈36% HF), PR-AUC is a more informative metric than ROC-AUC. The Precision–Recall plots (Fig. 10) show that XGB and RF outperform in the high-recall region, maintaining precision >0.9 up to recall ≈0.7. This demonstrates their ability to detect HF patients with minimal loss in precision, which is a critical requirement for clinical deployment where false alarms can be resource intensive.

**Figure 10:**
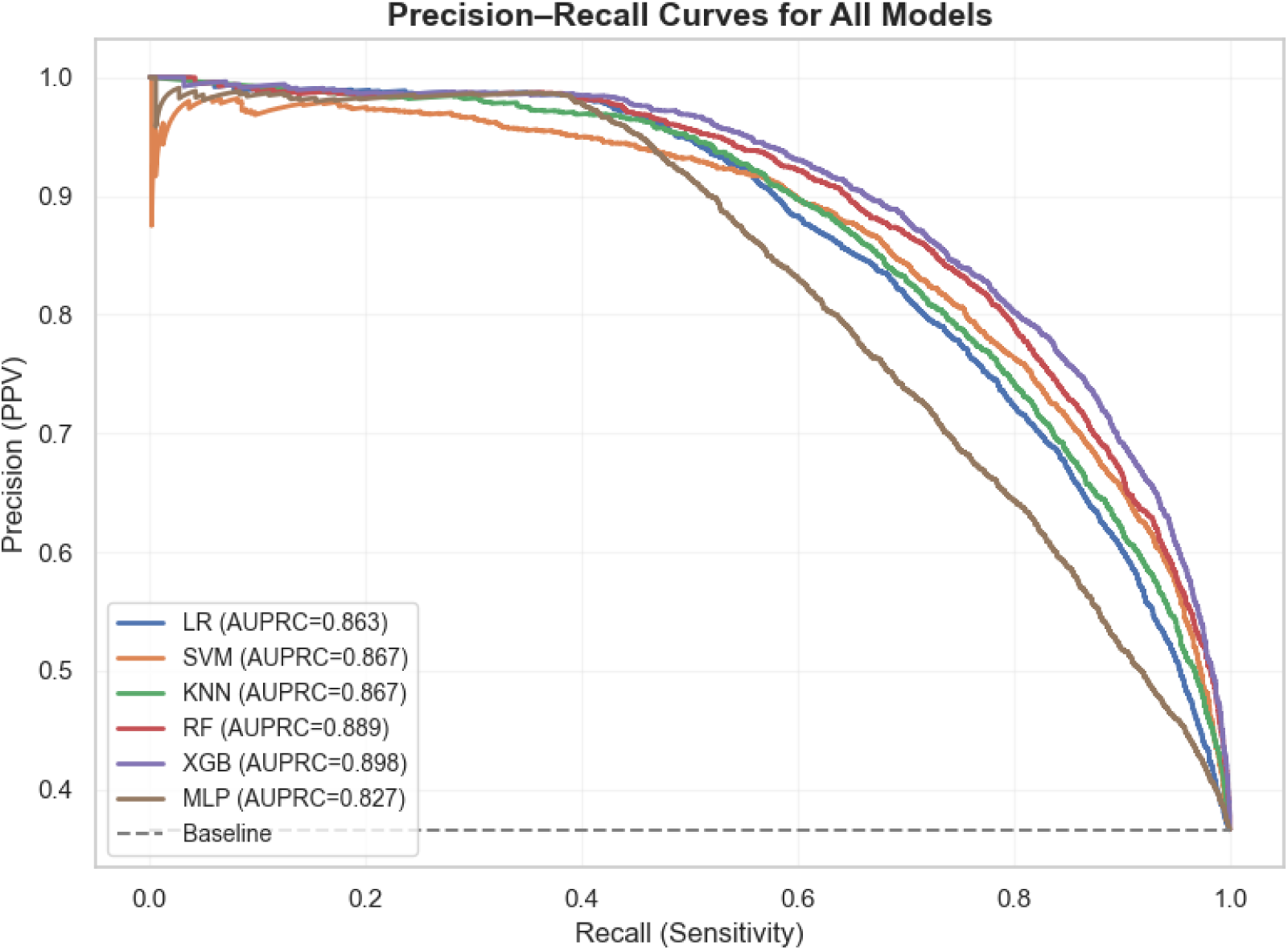
PR-AUC graphs for ML models

Further, confusion matrices were computed and plotted to quantify prediction distribution across classes (Fig. 11). XGB and RF models achieved the best balance, with true-positive HF detections (≈3,100–3,400) and low false-negative rates (≈650–990). The MLP overpredicted the majority class (non-HF), as seen from higher false negative counts, demonstrating sensitivity to class imbalance despite SMOTE implementation. SVM maintained competitive recall but generated slightly more false positives than tree-based models (RF and XGB).

**Figure 11:**
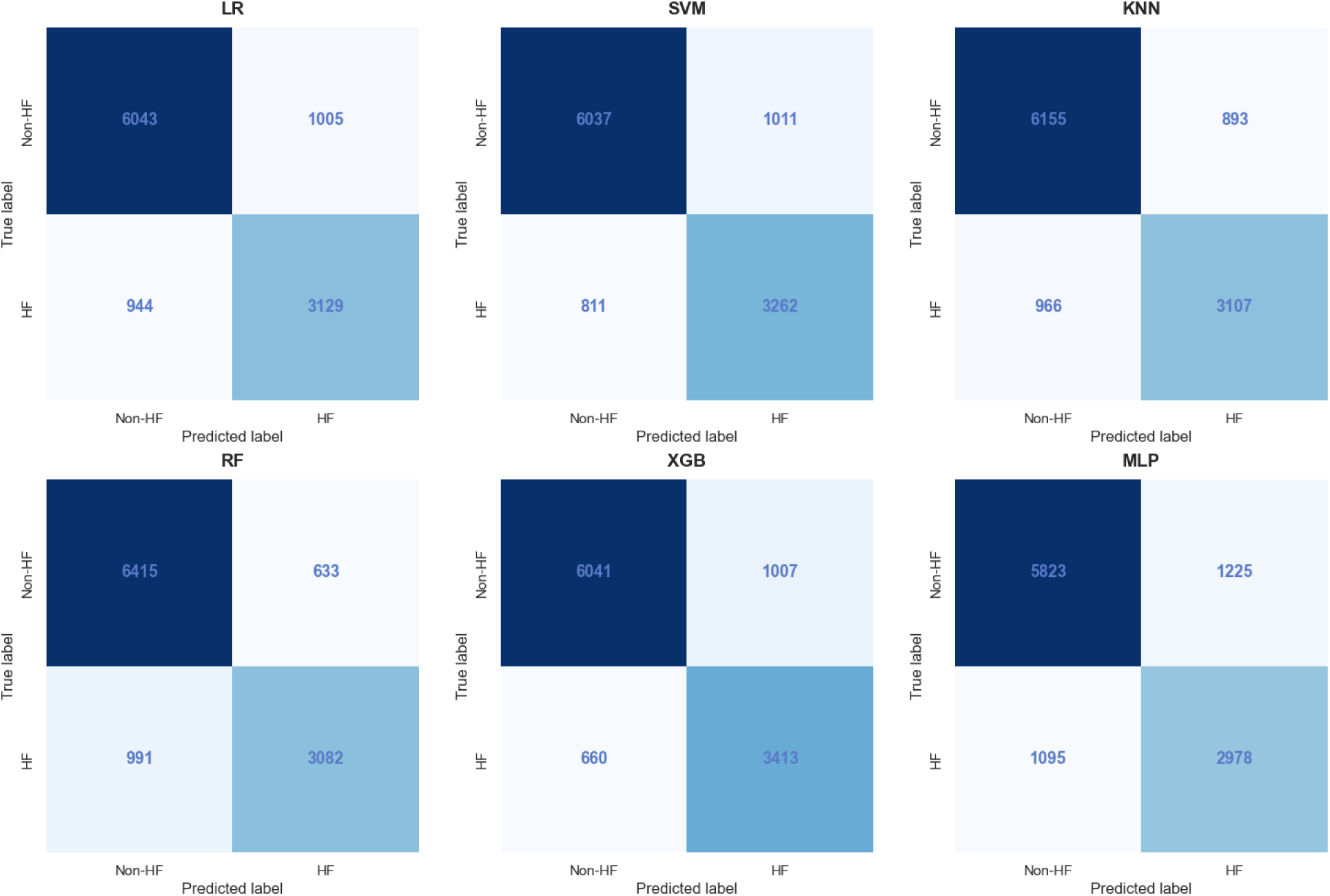
Confusion matrix for ML models

To understand how each feature influences the prediction we performed SHAP analysis. This analysis ranks features based on their impact and helps us to understand the rationale behind the AI model’s decision, which is essential aspect of explainable AI. Each dot shows one patient/sample, x-axis (SHAP value) represents the impact on prediction, and the color gradients signify the feature value (red – high value, blue – low value). The analysis (Fig.12) was derived from the XGB component of the stacked ensemble model; the top 10 most influential features included atrial fibrillation, age, hypertensive disorder, serum sodium, deprivation index, anemia, glucose, creatinine, hemoglobin, and blood pressure metrics ^39–42^. These results proved to be clinically consistent: atrial fibrillation and hypertension are well-established precursors of HF ^43^, while renal and hematologic abnormalities, including elevated creatinine and anemia, are well-established prognostic markers in heart failure ^44^. The color gradient shows that the higher values of sodium and deprivation index positively correlate with HF probability, while higher values of albumin, potassium, and HDL cholesterol lie on the negative x-axis, meaning elevated levels push prediction away from HF outcome, acting as a protective feature.

**Figure 12:**
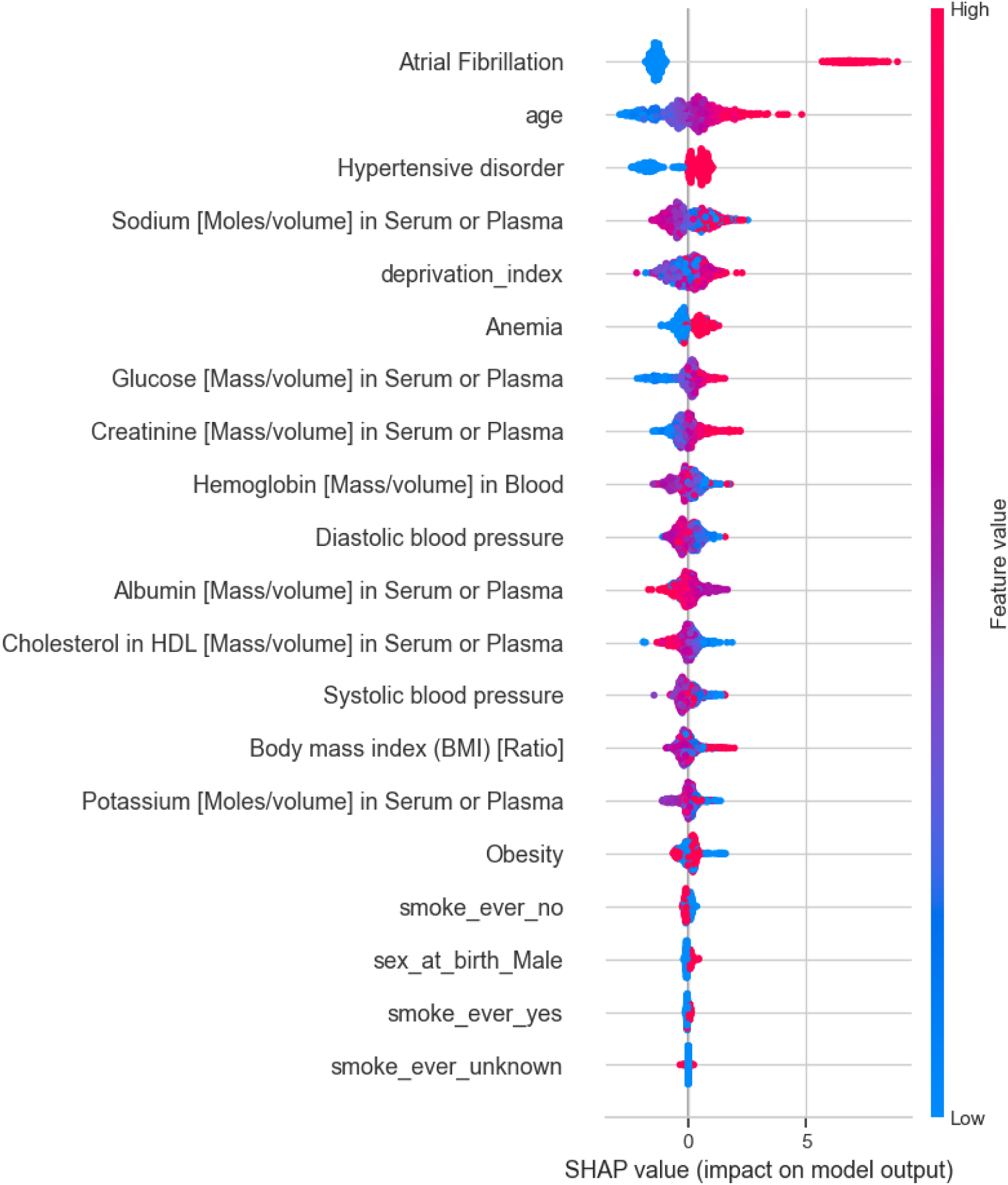
SHAP analysis extracted from the XGBoost component of the stacked ensemble model for explainability of feature importance and impact.

To validate our findings, we computed the feature correlation (Fig.13) which confirms SHAP findings of atrial fibrillation (r=0.53), age (r=0.39), and hypertensive disorder (r=0.34), proving to be the strongest positive correlators with HF incidence. Negative correlations of albumin, hemoglobin, and HDL align well with protective physiology. Whereas features like potassium and sodium demonstrate a minimal correlation with HF.

**Figure 13:**
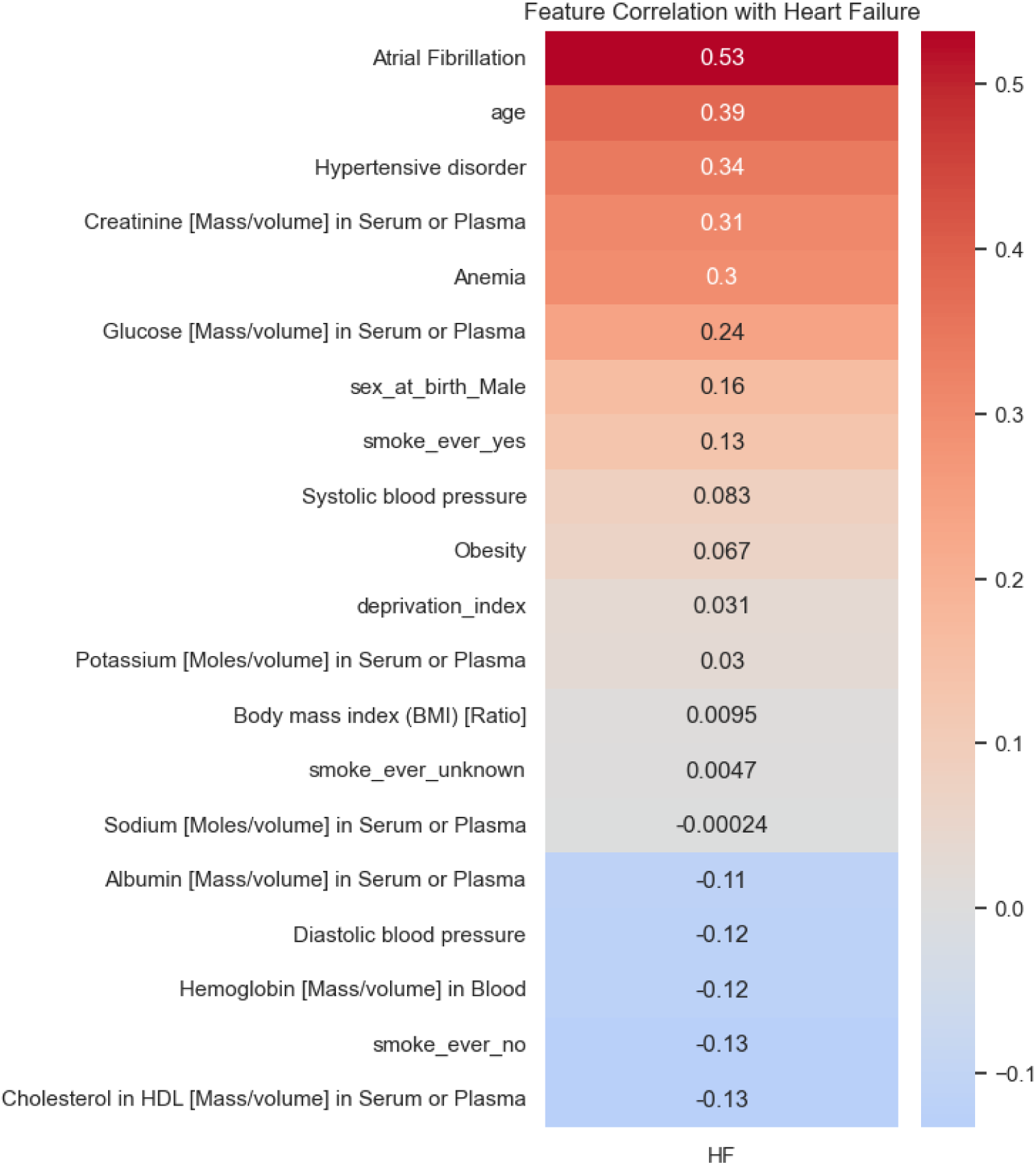
Feature correlation with HF..

## Secondary Experiment: Multiclass Heart Failure Subtype Classification

A secondary experiment was performed to attempt to classify the subcategories of HF; HFrEF and HFpEF. This prediction framework extended the previous binary classification problem to a multiclassification setting. The motivation for this experiment was to evaluate the model’s ability to differentiate among physiologically distinct HF subtypes, which is a more challenging and clinically relevant task ^45,46^. The same stacked ensemble model was trained on three settings (imbalanced, undersample, and oversample -SMOTE). This distributed the categories into No HF = 23,493, HFrEF = 5,426, and HFpEF = 3,831. Performance was assessed using precision, recall, F1 score, and macro-ROC-AUC (OVR). The results are compiled and shown in table 6 below:

**Table 6:** Stacked ensemble model performance on HF subtypes discrimination.

| SETUP<br>(TRAINING<br>STRATEGY) | CLASS | PRECISION | RECALL | F1-SCORE | AUC (OVR) | ACCURACY |
| --- | --- | --- | --- | --- | --- | --- |
| IMBALANCED<br>(NO<br>BALANCING) | No_HF | 0.856 | 0.944 | 0.898 | 0.8994 |  |
|  | HFrEF | 0.512 | 0.482 | 0.497 | 0.8630 |  |
|  | HFpEF | 0.452 | 0.151 | 0.227 | 0.8468 |  |
|  | **Macro Avg** | **0.607** | **0.526** | **0.540** | **0.870** | **0.795** |
|  | Weighted<br>Avg | 0.764 | 0.795 | 0.770 | — |  |
| UNDERSAMPLE<br>(FOLD-TRAIN<br>ONLY) | No_HF | 0.856 | 0.941 | 0.896 | 0.8955 |  |
|  | HFrEF | 0.519 | 0.459 | 0.487 | 0.8613 |  |
|  | HFpEF | 0.454 | 0.202 | 0.280 | 0.8432 |  |
|  | **Macro Avg** | **0.610** | **0.534** | **0.554** | **0.867** | **0.794** |
|  | Weighted<br>Avg | 0.765 | 0.794 | 0.773 | — |  |
| SMOTE (FOLD-TRAIN ONLY) | No_HF | 0.854 | 0.943 | 0.896 | 0.8953 |  |
|  | HFrEF | 0.499 | 0.447 | 0.471 | 0.8566 |  |
|  | HFpEF | 0.457 | 0.179 | 0.258 | 0.8411 |  |
|  | **Macro Avg** | **0.603** | **0.523** | **0.542** | **0.864** | **0.791** |
|  | Weighted<br>Avg | 0.761 | 0.791 | 0.768 | — |  |

The multiclass experiment revealed that the inherent difficulty in distinguishing specific HF subtypes might be due to overlapping clinical and laboratory features that do not directly encode the physiological distinctions that define these subtypes. In clinical practice, differentiation between the HF subtypes is determined by cardiac imaging (echocardiography) or other invasive imaging providing quantitative measures such as left ventricular ejection fraction and diastolic function. Although these measurements are considered the gold standard to differentiate HF subtypes, they require specialized equipment, trained personnel, and are difficult to perform in a resource-constrained environment. These imaging-based measurements were kept out of the scope of this study, as our main objective was to find whether low-cost, widely collected EMR features can support scalable HF identification.

With the unbalanced dataset, the model achieved the highest macro-ROC-AUC of 0.870, indicating strong overall discrimination. However, if we look into per-class recall, they were markedly lower for HFrEF (0.482) and HFpEF (0.151), demonstrating the dominance of the No HF majority class. This reflects the class imbalance impact commonly observed in clinical EHR data. Balancing by random undersampling improved minority recall for HFpEF (0.202), it lowered the macro-ROC-AUC (0.867). The reduction in available data limited the feature diversity. The SMOTE iteration generated an overall lower benchmark performance in comparison to others.

## Model Calibration and Risk Stratification

The stacked ensemble demonstrated strong discriminatory performance (ROC–AUC ≈ 0.9248; PR-AUC ≈ 0.8902), effectively ranking HF. However, discrimination does not guarantee that predicted probabilities represent clinically meaningful absolute risk. Model calibration is an important aspect, ensuring that a model’s predicted probabilities accurately reflect the likelihood of the outcomes and are considered crucial for reliable decision-making in clinical settings. Our analytic cohort consists of an HF prevalence of 36.6%, whereas typical real-world HF prevalence is substantially lower, typically around 1-3% in developed countries, according to data reported in 2023 ^2^. In such a cohort (36.6% prevalence), model probabilities may be numerically well calibrated but will have overestimated absolute risk when interpreted in the general population. Hence, for our stacked ensemble model, we performed a post-hoc probability calibration and then mapped it to the realistic base rate of 2.5% (an average estimation for current HF prevalence and considering the fact that each year the number of HF patients increases). It is important to note that we did not calculate the overall prevalence of HF using the formula: number of HF cases in the world / total population of the world (64 million cases worldwide in an 8 billion population), which gives us an approximate rate of 0.8%. This is because a base rate of 0.8% is likely an underestimate of the disease as HF is mostly underdiagnosed, and there is an incomplete global data as most of the regions rely on self-report.

### Cohort calibration and probability mapping

To calibrate our model, we applied Platt scaling method ^47^. To avoid test-label “peeking,” we subdivided our original training data into FIT subset (used to train) and a CAL subset (used to learn the calibration mapping), the held-out test set remained untouched throughout. Let *p̂_raw_* denote the raw predicted probability from the trained ensemble. Platt scaling fits a logistic recalibration on the log-odds:

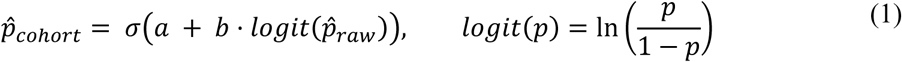

where *σ*() is the sigmoid function and parameters *a*, *b* are estimated on CAL subset. This systematically adjusts the confidence of probabilities while preserving the rank order. After this calibration the *p̂_cohort_* reflects the base rate of 36.6%. Hence, to map the probabilities to absolute risk scoring, we used the standard prior probability shift in log-odds space ^48^.

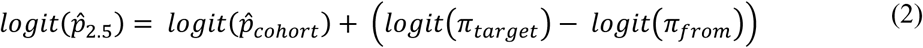

Where *π*_*target*_ = 0.025 (2.5% realistic prevalence rate) and *π*_*from*_ = 0.366 (36.6% cohort prevalence rate) from CAL subset. The resulting *p̂*_2.5_ can be interpreted as the model’s estimated probability of HF in a setting with a 2.5% prevalence rate. Since our test set still remains enriched by our cohort prevalence, we applied importance reweighting ^49^. The non-HF cases were assigned weight *w*_0_ = 1, and HF cases were assigned weight *w*_1_ such that the weighted prevalence equals 2.5% as shown in equation 3.

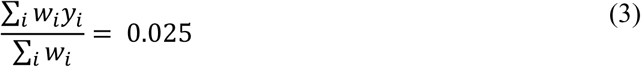

The importance reweighting produced an effective evaluation distribution matching the target base rate of 2.5% (weighted prevalence = 0.0250). The mapped probabilities were also aligned (weighted mean predicted risk = 0.02486), demonstrating model’s average predicted risk in sync with the expected event rate in the target population. To evaluate the accuracies of these probabilities, we calculated the weighted Brier score ^50^ and the weighted calibration intercept and slope to evaluate systematic bias and over/under confidence, following the established recommendations for clinical prediction models^51^. These results are summarized in table 7 with an explanation of what they represent and why they are important in our case.

**Table 7:** Calibration summary under 2.5% target prevalence. (Values computed on the held-out test set using importance reweighting to reflect a 2.5% deployment prevalence.)

| METRIC | VALUE | IDEAL / TARGET | WHAT IT REPRESENTS | WHY IT MATTERS CLINICALLY |
| --- | --- | --- | --- | --- |
| **TARGET PREVALENCE (Π_TARGET)** | 0.0250 | — | Assumed real-world HF prevalence for the deployment scenario | Defines the baseline risk environment in which absolute probabilities are interpreted |
| **WEIGHTED PREVALENCE** | 0.0250 | 0.0250 | Effective event rate after reweighting the enriched test set | Confirms evaluation reflects the intended real-world base rate rather than the enriched cohort |
| **WEIGHTED MEAN PREDICTED RISK** | 0.02486 | 0.0250 | Average predicted probability under the 2.5% scenario | Ensures probabilities are not systematically too high or too low at the population level |
| **WEIGHTED BRIER SCORE (2.5%)** | 0.01775 | 0 (lower is better) | Mean squared error between outcomes and predicted probabilities | Summarizes overall probabilistic accuracy of risk estimates |
| **WEIGHTED CALIBRATION INTERCEPT** | 0.01453 | 0 | Systematic bias in predicted log-odds (positive implies slight overestimation) | Detects global over- or underestimation of risk |
| **WEIGHTED CALIBRATION SLOPE** | 1.00268 | 1 | Over- or under-confidence of predictions | Ensures probabilities are neither exaggerated nor overly conservative |

Collectively, these metrics indicate that after these steps, the probability estimates are closer to the desired 2.5% HF prevalence population. In practical terms, if 1,000 patients were to be screened in a 2.5% prevalence setting, the expected number of HF cases is ∼25, and our model predicts ∼24.86 cases on average (0.02486 × 1000), demonstrating that the probability outputs are well-calibrated for absolute risk scoring in clinical deployment.

### Risk Stratification

After cohort calibration and prevalence rebasing, the model outputs probabilities *p̂*_2.5_ that can be interpreted as an individual’s estimated HF probability in a population where the overall HF prevalence is 2.5%. The next step for clinical usefulness is converting continuous probabilities into actionable groups to answer the question, “Who is at higher risk of having HF than whom?” or “Who is at highest risk of a HF diagnosis?” This is especially important in a resource-constrained setting where this model can support clinical decision-making, prioritizing follow-up for a smaller subset of patients while capturing a large fraction of true HF cases. To achieve that, instead of choosing one threshold, our model assigns each person a likelihood of having HF and splits them into 10 equal-sized groups (weighted risk deciles), from lowest to highest likelihood of having HF. The resulting decile cutoffs were 0.0024003, 0.0024586, 0.0025580, 0.0027517, 0.0030777, 0.0037576, 0.0054499, 0.0107362, and 0.0436975. Within each decile, we compared mean predicted risk to the weighted observed HF rate, where Decile 0 = lowest 10% predicted likelihood and Decile 9 = highest 10% predicted likelihood as shown in figure 14.

**Figure 14:**
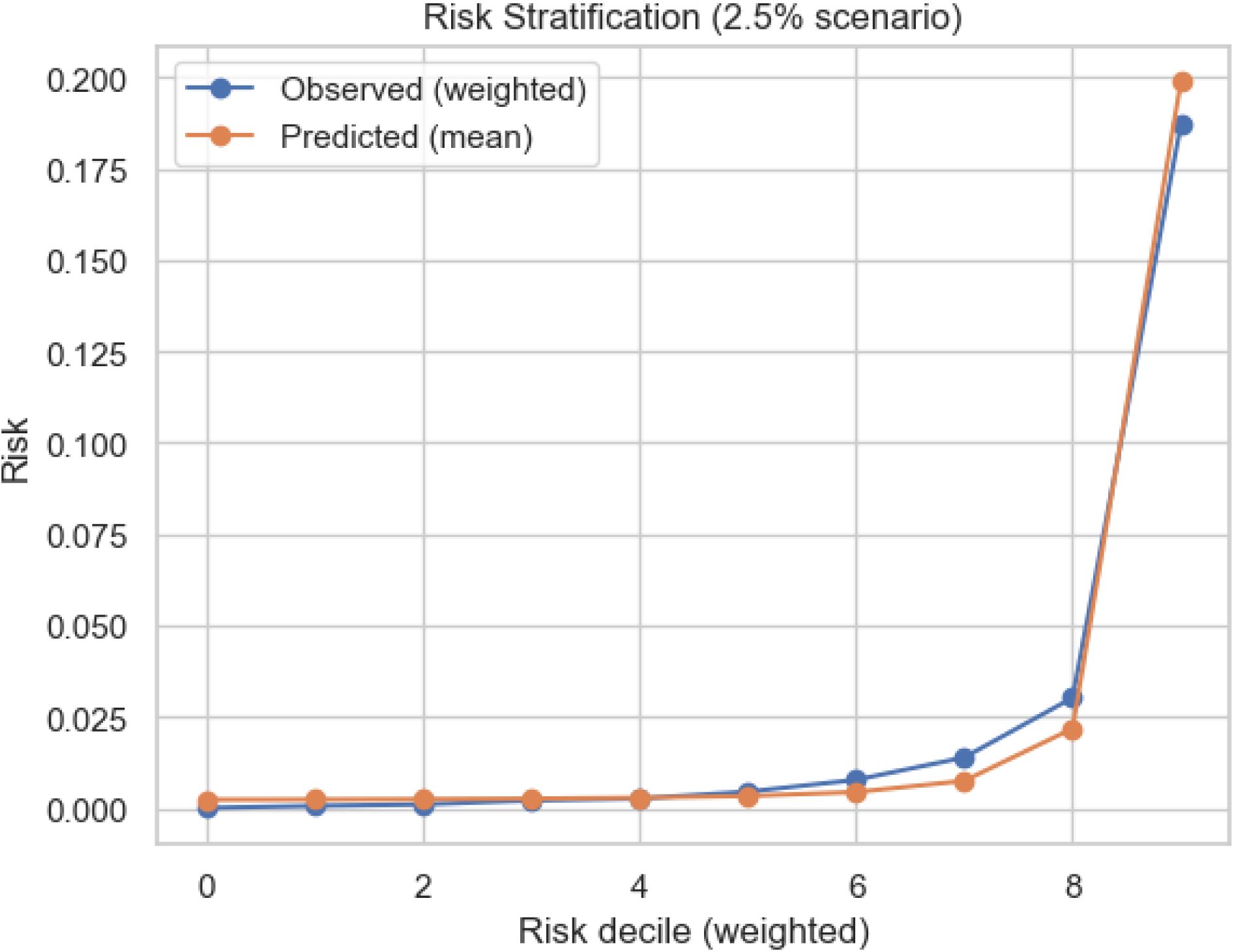
Predicted mean risk versus weighted observed HF rate across weighted risk deciles. Orange (Predicted mean): the average predicted probability *p̂*_2.5_ inside that decile. Blue (Observed weighted): the observed HF rate in that decile (but computed in a way that reflects a 2.5% population)

The plot shows that the observed predicted mean and average predicted probabilities closely follow each other, demonstrating a well-calibrated model. The lower deciles show almost zero likelihood of HF and then jump sharply as they approach the top decile.

To quantify the screening efficiency, we plotted a cumulative capture analysis. Patients were sorted from highest to lowest *p̂*_2.5_ and we computed the fractions of HF cases captured when selecting the top X% of the population. Results showed under the 2.5% scenario, the top 10% highest-risk individuals captured ∼74.7% of HF cases, the top 20% captured ∼86.8%, and the top 30% captured ∼92.4% as shown in figure 15. This directly answers a practical question: “If a clinic can only act on a limited fraction of patients, how many HF cases are still identified*?”* meaning if out of a population of 100,000 with an HF prevalence rate of 2.5% (2,500 cases of HF), a clinic screened the top 10% (10,000) ranked by this model, they would find 74.7% (1,868) of HF cases.

**Figure 15:**
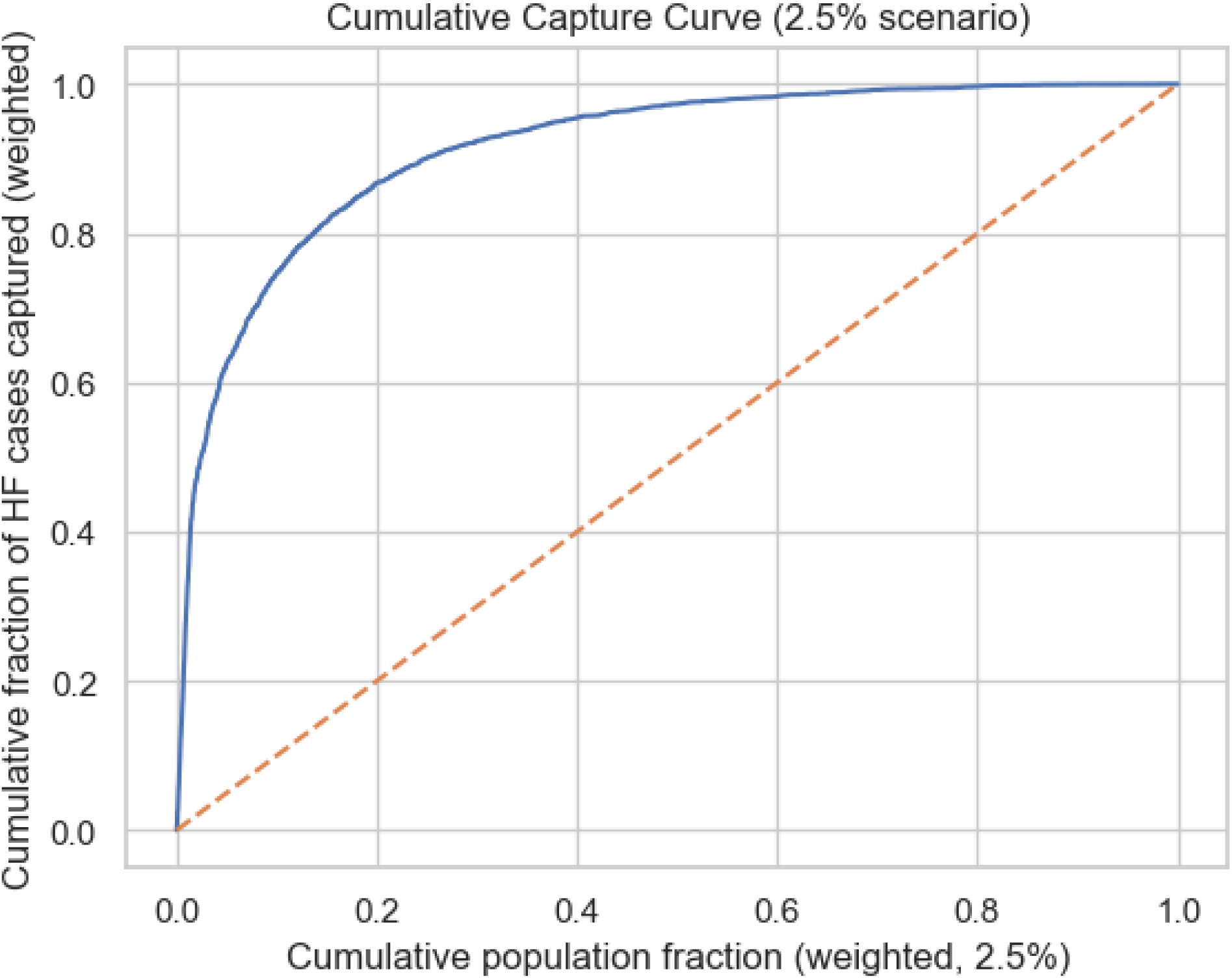
Cumulative HF cases captured vs the fraction of the population screened from highest to lowest risk.

## Discussion

This research developed and evaluated a comprehensive machine learning (ML) model framework for HF diagnosis and subtype classification of HF using large-scale electronic medical record (EMR) patient data from All of Us Research Program. The participants of the dataset consisted of 37,070 (13,577 HF patients, 23,493 non-HF controls) with further subgroups of 3 phenotypes: HFrEF (n = 5426), and HFpEF (n = 3831). In each record, there were 18 routinely available, inexpensive clinical and demographic variables (including measurements (e.g., albumin, creatinine, glucose, hemoglobin, blood pressure), comorbidity (e.g., atrial fibrillation, hypertension, obesity, anemia), lifestyle factors (e.g., smoking history), and demographics (age, sex, deprivation index).

The use of a multi-stage experimental pipeline (Fig. 7) involving data preprocessing (IQR-winsorization, median imputation, quantile transformer, one-hot encoding), class-imbalance management, model training, interpretability analysis, and risk stratification. The modeling was based on both the baseline classifiers, which included LR, SVM, KNN, Random Forest, XGBoost, and MLP as well as on a custom stacked ensemble with XGBoost, LightGBM, CatBoost, and MLP as base learners and a LR meta-learner. Three data-balancing methods were compared, random undersampling, SMOTE oversampling, and natural (unaltered) distribution. There were several complementary measures of model evaluation including ROC-AUC, PR-AUC, accuracy, precision, recall, and F1-score, followed by model calibration which helped to make sure that the model evaluation was robust even in the case of strong class imbalance.

It is essential to note that when comparing our predictive performance to prior ML studies, we must understand the prognostic vs. diagnostic nature of the research. Imaging-based works such as Matsumoto et al. ^17^ and Choi et al. ^5,18^ primarily focus on HF diagnosis/phenotyping using chest X-ray or echocardiography. Although they achieve strong accuracy, they require specialized, higher-cost modalities and operator expertise. On the other hand, EMR studies (Adler et al. ^20^, Ishaq et al. ^21^, and Wang ^22^) mainly focus on prognosis, often relying on temporal follow-up information. Hence, for these EMR studies we cannot do 1 to 1 comparison. In diagnostic prediction, Wu et al. ^19^ predicted HF up to 6 months before clinical diagnosis with AUC ≈ 0.77 but used under-sampling to balance classes, diverging the study from real-world prevalence and affecting probability interpretation. In contrast, our stacked ensemble scored considerably better in the binary HF-vs-non-HF diagnostics with ROC-AUC = 0.927, PR-AUC = 0.895, and accuracy = 0.856. SHAP analysis (Fig. 12) indicated that established HF pathophysiology (atrial fibrillation, hypertension, high creatinine, glucose, low hemoglobin, albumin, and higher deprivation index) was captured as a key predictor, demonstrating both biological coherence and clinical interpretability.

A second experiment with 3 diagnostic classes, namely No-HF, HFrEF, and HFpEF, with the same set of features and the same ensemble strategy was conducted. It had subpar performance, the best setup for this multiclassification was again achieved by using imbalanced setup (unaltered) dataset, obtaining a macro-ROC-AUC of 0.87 and an accuracy of 0.795. This is as expected of diagnostic complexity: HFpEF/HFrEF nomenclature may be substantially noisy in routine EMR due to the frequent requirement of echocardiography and standardized score systems like H2FPEF or HFA-PEFF, which combine imaging and natriuretic peptide and are inconsistently present in real-world data ^52–54^. The combination of these factors with the significant imbalance of classes and the overlap of clinical phenotypes probably dilutes the subtype discrimination in the case of the application of non-imaging, cross-sectional features alone. Although the minority-class recall was still limited because of the data imbalance, the experiment showed that EMR-only features have the potential to partially discriminate HF phenotypes without any imaging or temporal data, indicating that scalable HF subtyping with additional features could be made possible. Future work will focus on improving subtype modeling by (i) incorporating echocardiography features where available and (ii) validating subtype labels to reduce noise and misclassification, which may be a more clinically aligned and data-efficient pathway.

The analytic cohort used in this study had an HF with a ≈36.6% prevalence. Hence, we performed a post-hoc probability calibration and deployment-scenario adjustment to produce clinically interpretable absolute probabilities. Specifically, Platt scaling was implemented on a held-out calibration split to calibrate our model to the 36.6% prevalence rate of our cohort, followed by a log-odds prior adjustment to express predictions under a realistic 2.5% population prevalence. Under a 2.5% scenario, we obtained a well-calibrated results (weighted mean predicted risk 0.0249 vs. target 0.0250) with a weighted Brier score of 0.0178 and a near-ideal weighted calibration slope (1.0027) and intercept (0.0145).

In HF literature, risk stratification has two common meanings: (i) prognostic stratification among patients already diagnosed with HF (mortality or re-hospitalization risk) ^55–57^, and (ii) stratification of individuals by likelihood of HF presence or development of HF in near future ^58–61^, mostly relying on temporal data. However, our study introduces a new novel risk stratification model targeting a different clinical decision point: diagnostic support and population screening, identifying patients who are most likely to currently have HF using the routinely available, low cost EMR variables at a single instance. This is crucial for patient outcomes, as earlier detection means earlier treatment and longer survival. After calibration and prevalence rebasing, we can convert these continuous probabilities into actionable strata using weighted deciles, showing a strong concentration of HF cases among the highest-risk fraction of the population (the top 10% captured 74.7% of HF cases; top 20% captured 86.8%).

The paper acknowledges a key limitation of an absence of external validation on an independent dataset. Although we used stratified splits and cross-validation within our cohort to reduce overfitting and provide stable performance estimates, internal validation alone cannot fully confirm generalizability of the model to patients of different demographics, missingness patterns, or diagnostic code practices. Hence, an evaluation on at least one external health system is needed to reassess the calibration under real-world prevalence.

The findings suggest that in case of strong ML frameworks and ensemble approaches, it is possible to obtain highly discriminative and well-calibrated models with the help of non-invasive EMR data to support AI-CDSS. The paradigm combines predictive performance, interpretability, and risk stratification within one system that may enable early HF screening, prioritize diagnostic tests, and help in prevention plans on a population level. Future developments can include prospective validation and real-world evaluation of the model on different geographical based clinical settings, deployment of these predictive scores in a real time CDSS to evaluate its applicability in real world scenarios, and incubating more readily available features which are more closely correlated with the subtypes of HF to improve the subcategory classification.

## Conclusion

This study developed and rigorously evaluated a custom stacked ensemble model and several other traditional ML model frameworks for heart failure (HF) prediction and subtype classification using readily available, non-invasive clinical variables. Empirically, our stacked ensemble demonstrated ROC-AUC = 0.927 and PR-AUC = 0.895 on the test set. To ensure the predicted probabilities are interpretable under real-world conditions, we applied post-hoc probability calibration on a held-out calibration split and then adjusted the calibrated probabilities to reflect a realistic 2.5% population prevalence. The calibrated model was well aligned (weighted mean predicted risk closely matching the 2.5% target) and produced clinically meaningful stratification, with a strong concentration of HF cases in the highest-scored subgroup, making it useful for clinical HF screening in a resource-constrained setting. SHAP analysis found that atrial fibrillation, age, hypertensive disorder, sodium, socioeconomic deprivation, anemia, glycemia and renal indices (glucose, creatinine), hemoglobin, and blood pressure were among the most influential drivers-consistent with established HF pathophysiology and epidemiology, highlighting both the feasibility and current data limitations of subtype discrimination from structured EHR variables alone. These findings point towards the potential of AI-CDSS to assist in HF diagnosis, enabling proactive disease management and care strategies for cardiovascular health.

## Acknowledgements

We gratefully acknowledge All of Us participants for their contributions, without whom this research would not have been possible. We also thank the National Institutes of Health’s <u>All of Us Research Program</u> for making available the participant data [and/or samples and/or cohort] examined in this study.

## Data Availability

The data analyzed in this study come from the Controlled Tier of the *All of Us* Research Program. The Public Tier dataset is available online and can be accessed from https://www.researchallofus.org/data-tools/data-snapshots/. The Controlled Tier of the All of Us Research Program is not publicly available due to privacy protections and program policy. Access is restricted to approved researchers who complete the required training and data use agreements and can apply for approval at the *All of Us* Researcher Workbench by following instructions from https://www.researchallofus.org/register/.

## Author Contributions

S.A is the lead author, designed the methodology, performed cohort construction and the complete ML and risk stratification analysis, created figures/tables, and wrote the original manuscript draft. M.A.L provided clinical/heart-failure domain guidance to ensure medical appropriateness of the study, reviewed and edited the manuscript. W.A is the corresponding author, supervised the project, contributed to study design and scientific direction, and reviewed and edited the manuscript. All authors reviewed and approved the final manuscript.

## Code Availability

The underlying code for this study [and training/validation datasets] is not publicly available as all of the analysis is done on *All of Us* Researcher Workbench provided cloud Jupyter notebook platform. As per the policies researchers are not allowed to export any code files containing an explicit analysis of participant level data, except notebooks containing only aggregated results. The aggregated results notebook can be prepared and provided on request.

